# Comparison of feature-based indices derived from photoplethysmogram recorded from different body locations during lower body negative pressure

**DOI:** 10.1101/2025.05.02.25326908

**Authors:** Shrikant Chand, Neng-Tai Chiu, Yun-Hsin Chou, Aymen Alian, Kirk Shelley, Hau-Tieng Wu

**Affiliations:** Department of Mathematics, Duke University, Durham, NC, USA; School of Medicine, National Yang Ming Chiao Tung University, Taiwan; Department of Anesthesiology, Yale University, New Haven, CT, USA; Courant Institute of Mathematical Sciences, New York University, New York, NY, USA

**Author notes:** **Correspondence to:** Aymen Alian, Department of Anesthesiology, Yale School of Medicine, New Haven, Connecticut, United States of America., Hau-Tieng Wu, Department of Mathematics, Courant Institute of Mathematical Sciences, New York University, New York, New York, United States of America. These authors contribute equally to this work (Shrikant Chand, Neng-Tai Chiu). **Funding**: NONE. **Author’s contributions:** SC, NTC, YHC: data analysis and write-up. AA: idea, literature review, data collection, and write-up. KS: idea, literature review, data collection, and write-up. HTW: idea, literature review, data analysis, and write-up.

## Abstract

**Objective:** Various time domain features, including dicrotic notch (**dic**), in photoplethysmogram (PPG), and the pulse transit time (PTT) determined using the simultaneously recorded electrocardiogram (ECG), are believed to have a critical role with many potential clinical applications. However, the dependence of these parameters on PPG sensor location is less well known.

**Approach:** Three transmissive pulse oximetry probes (Xhale) were put simultaneously on the ear, nose, and finger of 36 healthy volunteers in the lower body negative pressure (LBNP) experiment. Various features of the recorded PPG signals were analyzed across different LBNP phases for each location. Simultaneously recorded finger PPG and ECG (Nellcor) were used to assess the dependence of PTT on PPG sensor location.

**Main Results:** PPG signal quality varies by measurement site, with nasal PPG showing the highest quality and ear PPG the lowest. Except pulse rate (PR), most feature-related indices differ across sites. Specifically, the ratios of detectable **dic** vary, highest in finger PPG and lowest in nasal PPG. When **dic** is detectable, the e point and **dic** are significantly different. Pulse rate variability indices and PTT also vary by location, though no clear conclusions can be drawn about PTT behavior across different LBNP phases.

**Significance:** Various indices derived from PPG signals in a well-controlled study environment are influenced by sensor placement. Although not all possible indices are examined, the findings clearly illustrate the sensitivity of signal features to measurement location. While these results may not be directly generalizable to routine clinical settings, caution is warranted when extrapolating findings from one PPG site to another. This consideration is especially important in the digital health era, where mobile devices with PPG sensors are increasingly deployed at diverse body sites.

## 1. Introduction

Photoplethysmography (PPG) is a non-invasive technique that measures relative blood volume changes in the blood vessels [1, 2]. In addition to its wide application in oxygen saturation evaluation, it contains rich information, like heart rate, heart rate variability, respiration [2, 3], venous oxygenation [4], sympathetic nervous system activity [5] and thermoregulation [6]. As an oscillatory signal, PPG contains a cardiac component that is attributed to hemodynamics. While a detailed review of PPG applications is beyond the scope of this study, prior work has shown that features of the cardiac component can provide clinically useful hemodynamic insights, such as detecting blood loss [7] or indicating early signs of central hypovolemia [8] [9] or estimating blood pressure [10].

A common approach for extracting hemodynamic information from PPG involves analyzing various features within cardiac cycles [11]. Commonly utilized features in a PPG cardiac cycle are outlined in Figure 1, where we follow the terminology used in Figure 4.13 in [12]. Key physiological landmarks of the PPG waveform include the pulse onset, systolic peak (**sys**), maximum slope point (**msp**), dicrotic notch (**dic**), and diastolic peak (if present). The systolic phase corresponds to the upslope initiated by ventricular contraction, while the downslope reflects the diastolic phase. We also compute area-based indices: A1 (between onset and **dic**) and A2 (between **dic** and the next onset), as illustrated in Figure 1. With these features, researchers design various indices to study hemodynamic status. A typical example is the reflective index (RI), which is defined as the ratio of the heights of **sys** and **dia** [13, 14, 46, 47]. RI is a measure of pulse wave reflection and the small vessel tone, and it is a more reliable index of the effect of vasoactive drugs. Another example is the stiffness index (SI), which is defined by the ratio of the subject height and the time delay between **sys** and **dia** [15, 16, 46, 47]. SI is used as a marker of large artery stiffness and vascular aging, and it is strongly correlated with the central pulse wave velocity. Moreover, its relationship with the mean pulmonary artery pressures and coronary heart disease have been reported in [17, 18], among others. When electrocardiogram (ECG) is available, pulse transit time (PTT) is another widely applied landmark-based feature derived from the time delay between the R peak in ECG and **msp** in the PPG. PTT is approximately inversely proportional to blood pressure and has been used as a non-invasive proxy for blood pressure estimation [19, 48]. More applications can be found in, for example [20]. Features-related indices considered in this paper are summarized in Table 1.

**Figure 1:**
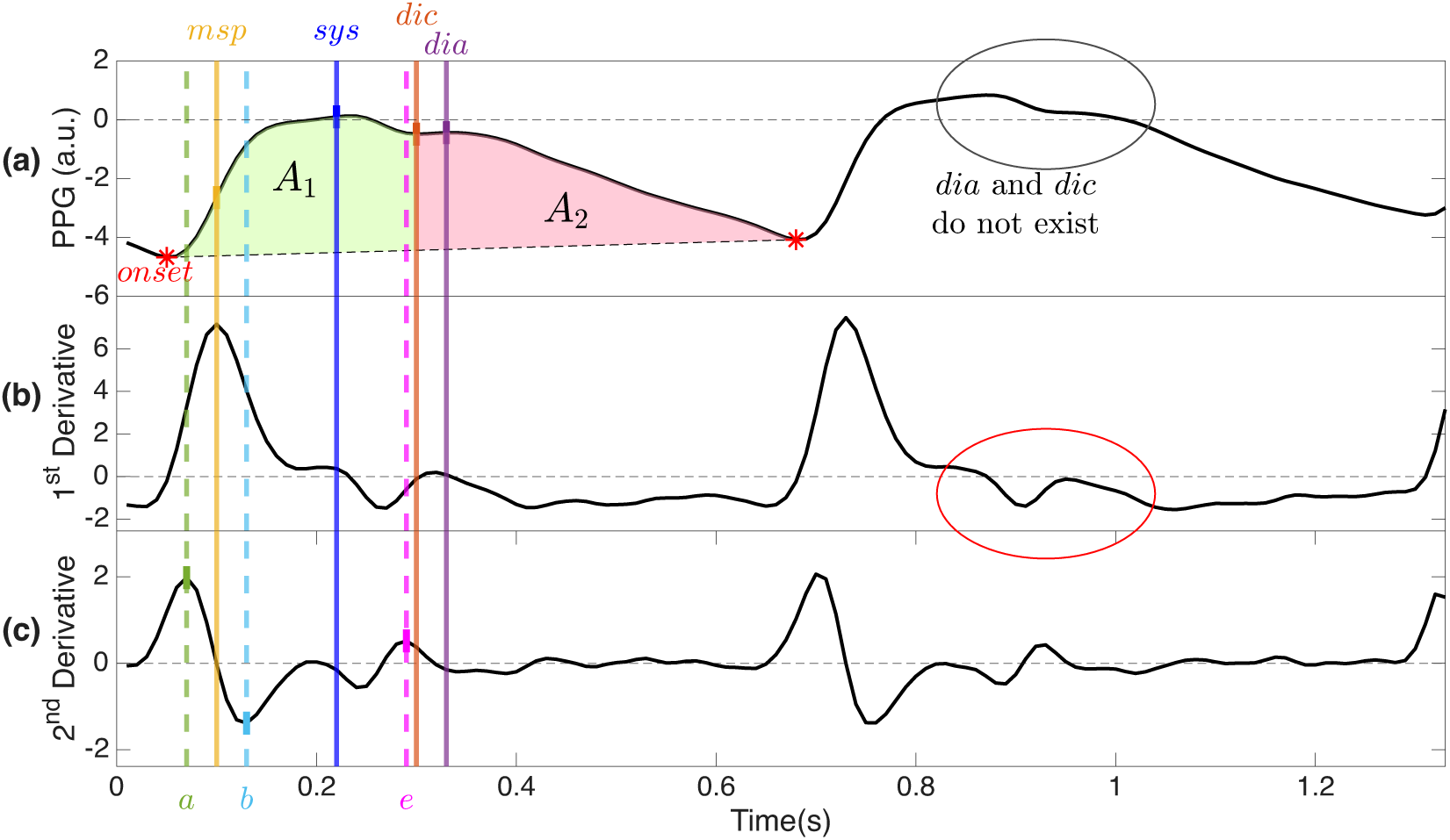
A summary of various landmarks. **(a)** an ear PPG signal (shown in black) as an example. The timestamp corresponding to the maximum of the first derivative of the PPG signal during the upper stroke of a cardiac cycle is denoted as **msp** (maximum slope point, shown in yellow). The peak situated between the upslope and downslope is termed the **sys**-peak (systolic peak, shown in dark blue), with the upslope representing the systolic phase, initiated by ventricular pressure transmission, and the downslope signifying the diastolic phase, associated with the slowdown of blood ejection from the ventricles. The local minimum between **sys** and the previous **sys** is called the pulse onset (**onset**, shown in red star). The peak in the downslope with a deflection, if exists, is called the ***dia****-peak* (diastolic peak, shown in purple), and the deflection point (or notch) is called the *dicrotic notch (**dic***, shown in orange). The area under the curve is also informative. The area under the curve (and above the line interpolated between the *i*^*th*^ and the next **onset**’s) between **onset** and **dic** (**dic** and the next **onset** respectively) is called A1 (A2 respective) and shown as the green (pink respectively) area. **(b)** The first derivative of ear PPG signal in (a), **(c)** the second derivative, sometimes called APPG, of ear PPG signal in (a) with a, b, and e points marked by the intersection of the signal with the green, light blue, and pink dashed vertical lines respectively. In this example, the second cardiac cycle does not have **dia** and **dic**, which can be observed that its first derivative is constantly negative around the region, marked by the red circle, that **dia** and **dic** should appear. a.u. means arbitrary unit.

**Table 1:** Feature related indices and their definitions, where *f* is the PPG signal. *δ*(·) is 1 if the condition inside is true, and 0 otherwise.

| Feature-related Index | Description |
| --- | --- |
| $PPT_i = t_i^{dia} - t_i^{sys}$ | **sys** to **dia** time. |
| $T2P_i = t_i^{sys} - t_i^{msp}$ | **msp** to **sys** time. |
| $RI_i = \frac{f(t_i^{dia})}{f(t_i^{sys})}$ | **dia** to **sys** height ratio. |
| $TT_i = t_i^{msp} - t_{i-1}^{msp}$ | Total cycle time. Only values less than 1.5 will be analyzed to avoid outliers. |
| $DNB_{i_m} = t_{i_m}^{msp} - t_{i_{m-1}}^{dic}$ | **dic** to baseline time. It is only evaluated when there is a detectable **dic**. Only values less than 1 will be analyzed to avoid outliers. |
| $A2A1_i = \frac{\int_{t_{i_{m-1}}^{dic}}^{t_{i_m}^{msp}} (f(s) - g(s))ds}{\int_{t_{i_m}^{msp}}^{t_{i_m}^{dic}} (f(s) - g(s))ds},$<br>where<br>$g(s) = \frac{f(t_{i_m}^{msp}) - f(t_{i_{m-1}}^{msp})}{t_{i_m}^{msp} - t_{i_{m-1}}^{msp}} \times (s - t_{i_m}^{msp}) + f(t_{i_m}^{msp})$ | Area 2 to Area 1 ratio, where Area 1 and Area 2 are areas between signal and interpolated line between two consecutive onsets. It is only evaluated when there is a detectable DN. Only those cycles with $t_{i_m}^{msp} - t_{i_{m-1}}^{msp}$ between **0.25 s** and **1.5 s** will be analyzed to avoid outliers. |
| $RMSSD_j = \sqrt{\frac{1}{n} \sum_{i=j}^{j+n} (PPI_i^{Rh} - PPI_{i-1}^{Rh})^2}$ | Root mean square of $n$ successive pulse to pulse interval differences in 5 minutes segment. $h=0$ means we use **msp** and $h=1$ means we use **sys**. The unit is milliseconds. |
| $SDNN_j = \sqrt{\frac{1}{n} \sum_{i=j}^{j+n} (PPI_i^{Rh} - \overline{PPI^{Rh}})^2}$ | Standard deviation of $n$ successive pulse to pulse interval over cycles in 5 minutes segment, where $\overline{PPI^{Rh}}$ is the mean. $h=0$ means we use **msp** and $h=1$ means we use **sys**. The unit is milliseconds. |
| $SDSD_j = \frac{1}{n} \sum_{i=j}^{j+n} (\Delta PPI_i^{R_h} - \overline{\Delta PPI^{R_h}})^2,$<br>where $\Delta PPI_i^{R_h} = PPI_i^{R_h} - PPI_{i-1}^{R_h}$ | Standard deviation of successive differences in $n$ pulse-to-pulse intervals over 5 minutes segment, where $\overline{\Delta PPI^{R_h}}$ is the mean of those differences. $h=0$ means we use **m<sub>sp</sub>** and $h=1$ means we use **s<sub>ys</sub>**. The unit is milliseconds. |
| $SD1_j = \sqrt{\frac{1}{2} SDSD_j}$ | Standard deviation perpendicular to the line-of-identity (the short axis of the Poincaré ellipse). The unit is milliseconds. |
| $SD2_j = \sqrt{2SDNN_j^2 - \frac{1}{2} SDSD_j}$ | Standard deviation along the line-of-identity (the long axis of the ellipse). The unit is milliseconds. |
| $pNN50_j = \frac{1}{n} \sum_{i=j}^{j+n} \delta(\vert PPI_i^{R_h} - PPI_{i-1}^{R_h}\vert > 50) \times 100$ | Proportion of successive pulse-to-pulse interval differences greater than 50 milliseconds, computed over 5 minutes segment. |
| $pNN20_j = \frac{1}{n} \sum_{i=j}^{j+n} \delta(\vert PPI_i^{R_h} - PPI_{i-1}^{R_h}\vert > 20) \times 100$ | Proportion of successive pulse-to-pulse interval differences greater than 20 milliseconds, computed over 5 minutes segment. |
| $LF_j$ | Power in the low frequency band (0.04 to 0.15 Hz) of the $PPI^{R_h}$ spectrum over 5 minutes. |
| $HF_j$ | Power in the high frequency band (0.15 to 0.4 Hz) of the $PPI^{R_h}$ spectrum over 5 minutes. |
| $LHR_j = LF_j / HF_j$ | The ratio of low and high frequency powers. |
| $PTT_j = t_j^{msp} - t_j^R$ | The time difference between the R peak ($t_j^R$) in the simultaneously recorded ECG signal and **m<sub>sp</sub>** of the associated cardiac cycle in the PPG signal. |

Although the feature-based approach is common, ample evidence suggests varying behaviors in PPG signals from different body sites [8, 21–24, 47, 50, 51]. This raises concerns about the compatibility of indices derived from diverse PPG signal sources, particularly with the proliferation of PPG-based mobile devices on different body locations [25]. A natural question regards the consistency or comparability of a feature-related index across diverse body locations where PPG signals are recorded. For example, certain features, such as **dia** and **dic**, may not always be present, as discussed in arterial blood pressure literature [7]. In such cases, a common approach involves using the e point derived from the second-order differentiation of PPG as a substitute. However, as **dic** may appear in one cycle and vanish in the next (see Figure 1 for an example, where **dic** is different from the e point), the accuracy of the e point’s positioning as a **dic** surrogate is crucial to avoid potential errors. This leads to another natural question: how different the **dic** and the e point are in different body locations. Another relevant topic is the potential existence of multiple local maxima around **sys**, complicating the determination of **sys** and consequently affecting indices reliant on it. See Figure 2 for an example. Given that **sys** and **msp** are typical features used to assess pulse rate (PR) and pulse rate variability (PRV), another natural question is about the variance in PR and PRV indices evaluated using **msp** or **sys** across different body locations.

**Figure 2:**
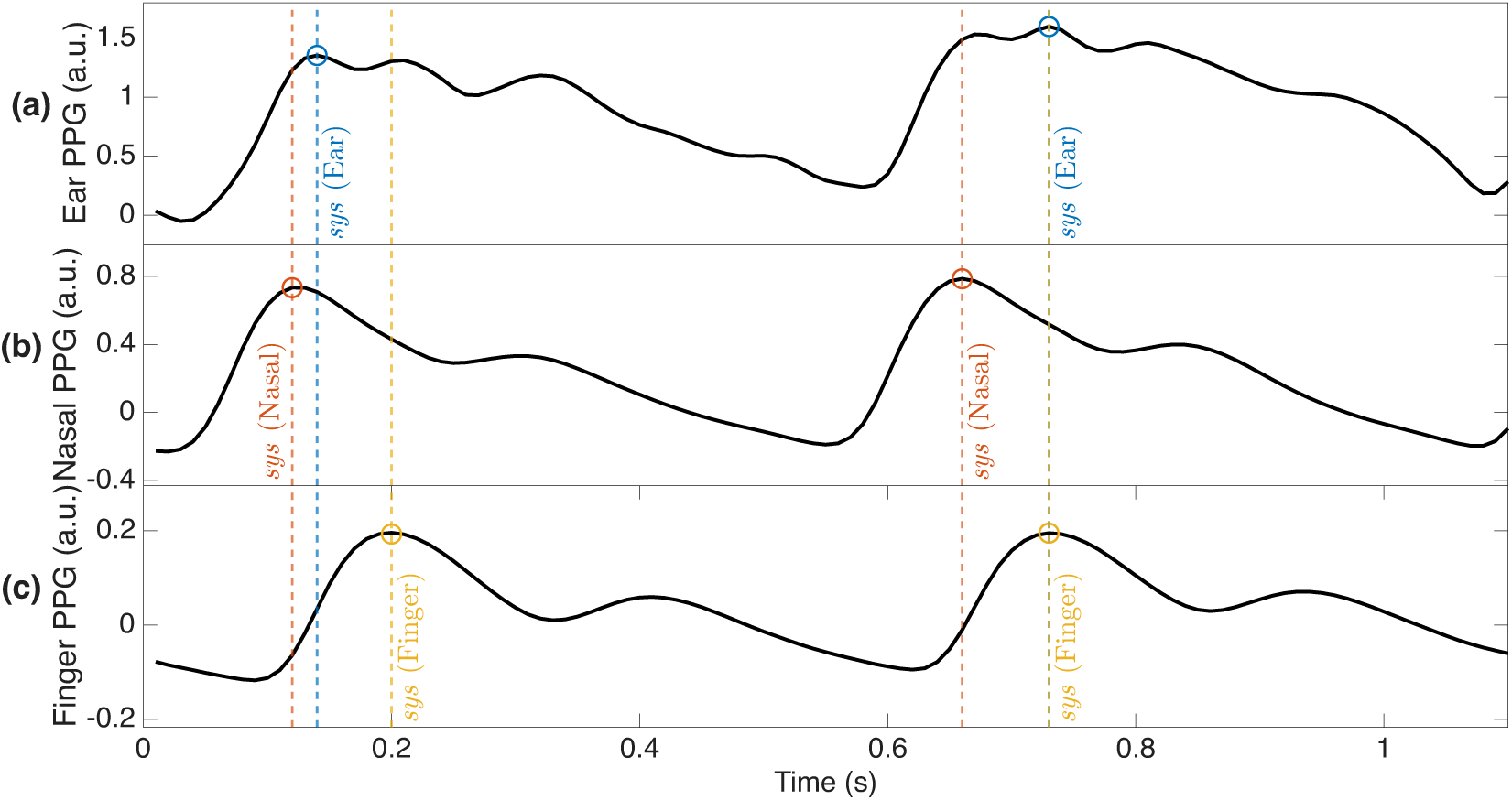
PPG signals with two cardiac cycles recorded from different body locations of a subject. The **sys** marked by the dashed lines are the **sys**’s detected from the ear PPG shown in (a). In the ear PPG, each cycle shows two local maxima around the locations where **sys** should exist. Notice that the first **sys** is detected at the first local maxima while the second **sys** is detected at the second local maxima. On the other hand, there is only one local maximum in the nasal PPG and finger PPG shown in (b) and (c) in this example. By visual inspection, clearly the location of **sys** for PPGs recorded from different body locations do not align. a.u. means arbitrary unit.

In this study, we investigate these questions through PPG signals simultaneously recorded from various body sites, such as the ear, nose, and finger, during a lower body negative pressure (LBNP) experiment through an integrated data acquisition system and consistent sensors from the same manufacturer. Based on existing literature and evidence, we hypothesize that even when using the same sensor, indices derived from PPG signals recorded at different body locations, ranging from those listed in Table 1 to phase harmonic index (PHI) and amplitude harmonic index (AHI) [8], are not necessarily equivalent. Since PTT reflects vascular path length and local arterial compliance, we also expect PTT to differ across sensor locations.

## 2. Material

We use a database collected from the LBNP study carried out in Yale-New Haven hospital. All procedures performed in studies involving human participants were in accordance with the ethical standards of the institutional and/or national research committee and written consent was provided. The research was conducted in accordance with the principles embodied in the Declaration of Helsinki. The Human Investigation Committee of the Yale School of Medicine approved the study protocol 2000020057. The clinical trial number is NCT 03592290. There are 36 healthy subjects (19 males and 17 females) with ages 18-40 recruited for the study. Exclusion criteria included current pregnancy or any history of pulmonary, cardiac, or vascular disease, including hypertension. A negative pregnancy test was confirmed for all female subjects. Subjects were instructed to refrain from consuming alcohol and caffeinated beverages, strenuous exercise, and tobacco consumption for at least 12 hours before the study. We focused on the Xhale transmissive SpO_2_ sensors (Xhale, Inc., Gainesville, FL) that were used to monitor PPG waveforms at the ear, nasal, and finger using identical sensors, and the Nellcor transmissive SpO_2_ sensor at the finger along with the ECG sensor. All data was digitized at 100Hz and continuously recorded to a laptop computer using a data acquisition system (LabChart, ADInstruments, Version 7.2.2). Specifics regarding the setup of the LBNP apparatus and the study environment have been previously described in detail [26] and we refer readers with interest there.

## 3. Methods and statistics

We start with detailing the algorithm that automatically annotates features of interest. Assume that the PPG is sampled uniformly with the period Δ_*t*_> 0 second (the sampling rate is 1/Δ_*t*_ Hz). The signal is mean-centered. Since various indices are with unit ms, the PPG signal is then upsampled to 500 Hz to increase the resolution. Subsequently, a high-pass filter with a cutoff frequency of 0.5 Hz and a low-pass filter at 20 Hz are applied. Broken segments containing obvious visual artifacts, such as flat lines or pure noise, lasting longer than one minute were excluded from the analysis. The resulting signal is saved as a vector *f*. Since our focus is the features of cardiac component, we consider high-quality PPG segments in *f* selected using the signal quality index (SQI), which was designed to select segments with high quality cardiac cycles. The SQI is outlined below. The PPG signals are first split into non-overlapping 5 second segments. The PPG quality for each segment is determined by the same ratio-based algorithm used in [27]. In short, using time-frequency analysis tools, the cardiac component of the PPG signal is decomposed into its strongest five harmonics and the remaining residue. The SQI is then set to be the ratio of the strength of these harmonic components to the total signal strength, which is a value between 0 and 1. The higher the score is, the better the PPG quality is. See Figure 3 for an example of PPG segments with different SQIs. We reject segments with a score lower than 0.9 and focus on the remaining segments, which we view as high-quality, for the following analysis. We choose 0.9 as the threshold based on our previous experience for feature analysis and visual inspection that segments with SQI greater than 0.9 usually have identifiable cardiac cycles and desired features.

**Figure 3:**
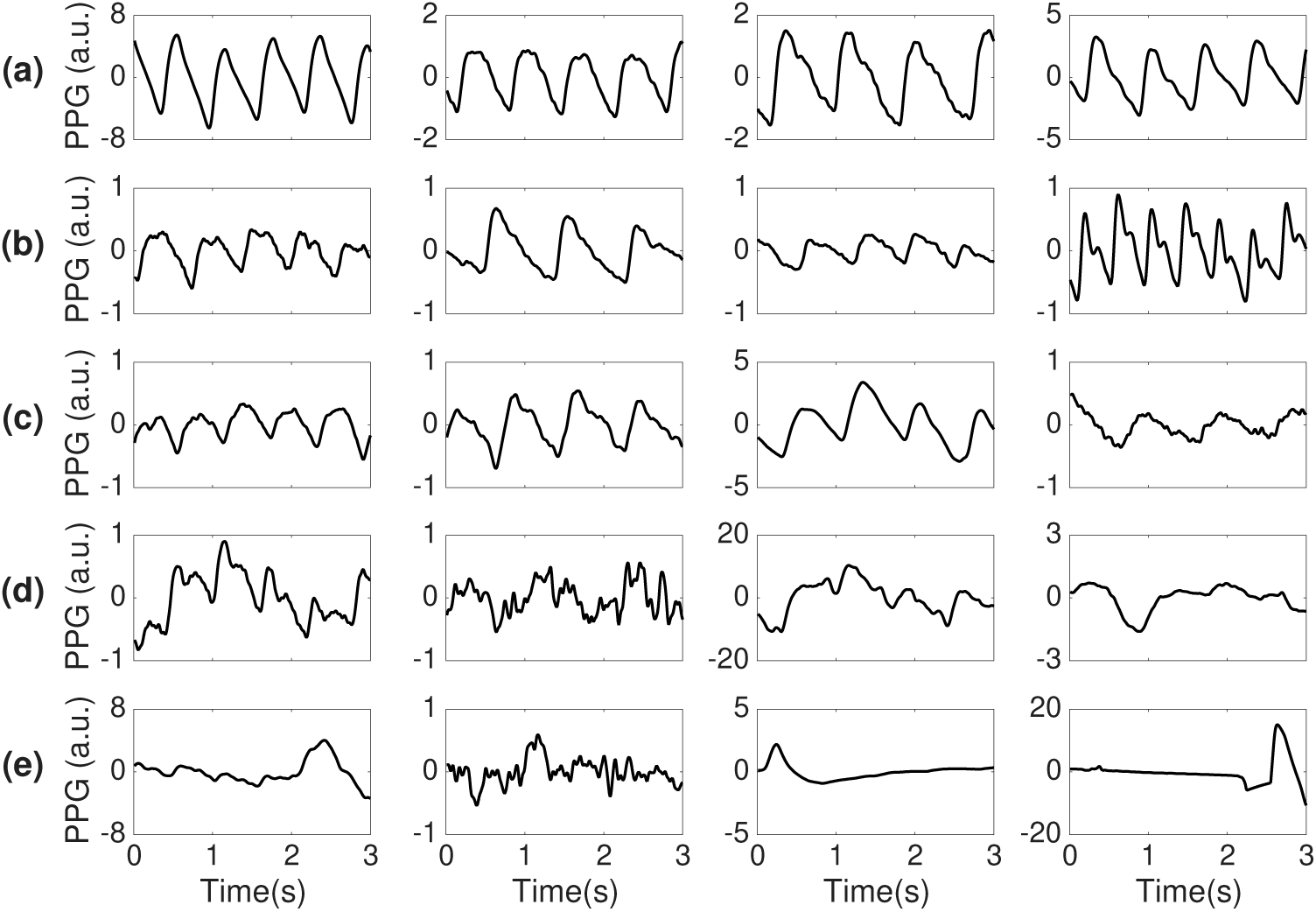
Ear PPG signals with an SQI of **(a)** 0.99, **(b)** 0.95, **(c)** 0.9, (d) 0.5, (e) 0.1. a.u. means arbitrary unit.

**sys** is determined as the local maximum of each cardiac cycle in the PPG signal using the standard double kernel technique. For a given cardiac cycle, **onset** is determined as the local minimum between the previous **sys** and the current **sys**. **msp** is determined from the local maximum between **onset** and **sys** in the first derivative of PPG. It is important to mention that while any numerical differentiation method may be considered [28], to be reproducible, we consistently evaluate the second derivative by carrying out two iterations of the centered finite difference method at interior points and two iterations of the forward/backward finite difference method at boundary points so that the accuracy of numerical evaluation is of order (Δ_*t*_)^2^ and Δ_*t*_in the interior and boundary points respectively. **dia** is then determined as the local maxima between **sys** and **onset** of the next cardiac cycle, if exists. Note that **dia** may not always exist. Denote 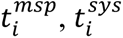 and 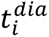 to be the time stamps of **msp**, **sys**, and **dia**, respectively, of the *i*-th cardiac cycle (see **Figure 1**).

When **dia** exists, we could determine **dic** as the local minimum between **sys** and **dia**, which is determined in the following way. Evaluate the second derivative of PPG, which we call the *acceleration PPG* (APPG). We look for APPG’s first local maximum after the detected **sys**. We label this point the e point, which again we follow the notation shown in (Figure 4.13 in [12]). Once the e point is detected, we look back to the original PPG signal for the local minimum in a small interval of 1/40 seconds around the e point. If it exists, we label this the **dic** and say that this cardiac cycle has a *detectable **dic***; otherwise, we call this cardiac cycle ***dic****-free*. Suppose there are *N* cardiac cycles in the high-quality PPG signal and there are *M* ≤ *N* cycles that have detectable **dic** in the time domain, indexed as *i*_1_ < ⋯ < *i*_*M*_, and the timestamps of these **dic**’s are denoted as 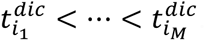. Also denote the timestamps of the e-point of all N cycles as 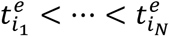 (see **Figure 1**).

**Figure 4:**
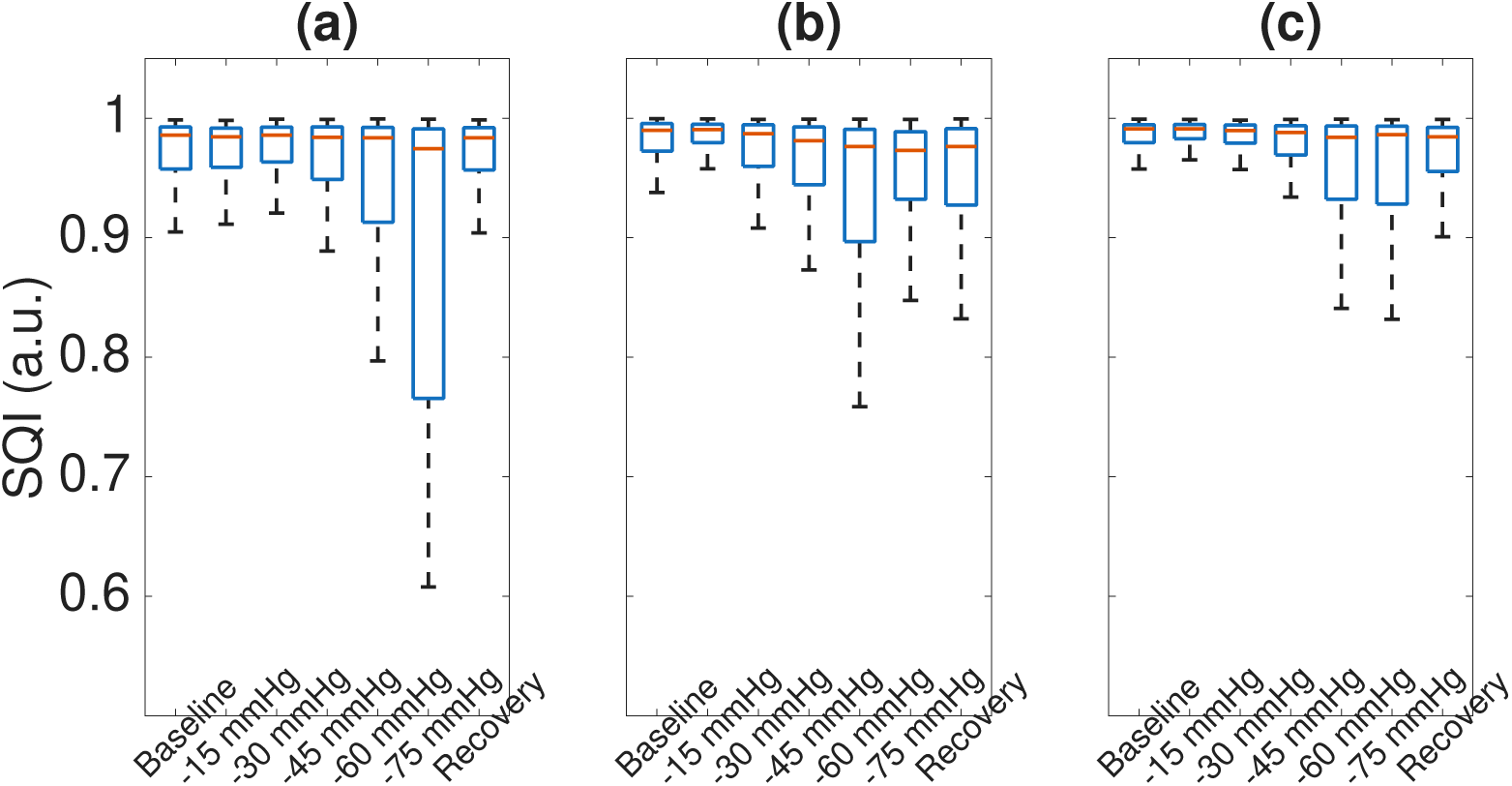
Progression of the distribution of signal quality index (SQI) over various experimental stages for **(a)** finger PPG, **(b)** ear PPG, and **(c)** nasal PPG.

With the above features, we consider the following feature-based indices. For *i* = *i*_1_, … , *i*_*N*_, define 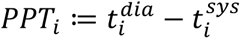 and 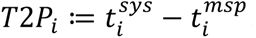. The reflective index (RI) is defined as 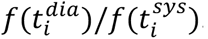. For *i* = *i*_1_, … , *i*_*N*–1_, the total cycle time 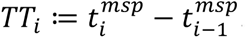. For *i* = *i*_1_, … , *i*_*M*_, we take the **dic** to baseline time (DNB) as 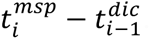. We also evaluate the A2 to A1 ratio, called A2A1. See Table 1 for a summary of these indices.

To evaluate PR and PRV, we consider **msp** or **sys** as the surrogate of R peak in the ECG signal. The PR is defined as the mean of the inverse of consecutive **msp**-**msp** or **sys**-**sys** intervals over 25 consecutive cardiac cycles, which we denote as **msp**-PR or **sys**-PR. Define the interval between two consecutive **msp**’s or **sys**’s as the pulse-to-pulse intervals (PPI), which is a surrogate of the R-peak to R-peak interval (RRI) when we study heart rate variability (HRV) in ECG signals [29]. Using PPIs, we evaluate nine PRV indices, including the root mean square of successive differences between normal heartbeats (RMSSD), the standard deviation of all normal RR intervals (SDNN), Poincaré plot-derived measures (SD1 and SD2), the proportion of successive differences exceeding 50 ms and 20 ms (pNN50 and pNN20), and frequency-domain features such as low-frequency power (LF, 0.04-0.15 Hz), high-frequency power (HF, 0.15-0.40 Hz), and their ratio (LHR=LF/HF). If **msp** is used to define PPIs, we call these indices **msp**-RMSSD, **msp**-SDNN, etc.; otherwise, we call them **sys**-RMSSD, **sys**-SDNN, etc. These indices are computed over 5-minute epochs with 4 minutes overlap from the high-quality segments, and RRI shorter than 0.25 s or longer than 1.5 s are excluded from the time-domain indices calculation.

To quantify phase and amplitude behaviors of the harmonics of a signal, we collect AHI and PHI indices which are summarized from [8]. Following the adaptive harmonic model (AHM), we suppose the PPG signal expresses a decomposition of the form

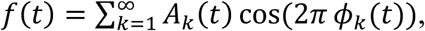

where *A*_*k*_(*t*) and *ϕ*_*k*_(*t*) are the time-varying amplitude and phase, respectively, of the

*k*-th harmonic component at time *t*. To quantify the phase behavior over a time interval *I*, we define the *k*-th phase hemodynamic index (PHI) as

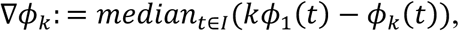

which is the phase discrepancy between the *k*-th harmonic and the fundamental component, where *k* = 2,3. Similarly, we define the *k*-th amplitude hemodynamic index (AHI) as

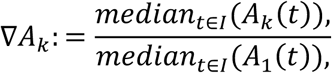

which is the phase discrepancy between the *k*-th harmonic and the fundamental component, where *k* = 2,3.

Finally, we examine how PTT varies with PPG sensor location. Given that ECG and PPG signals were collected from different instruments, we first aligned them using the following approach. We construct the instantaneous pulse rate (IPR) from the **msp** of the PPG, denoted as **msp**-IPR, by applying cubic spline interpolation to the nonuniformly sampled data points 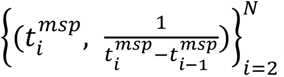, where *N* represents the number of cardiac cycles. Signal alignment between Nellcor and Xhale is achieved by maximizing the correlation between the **msp**-IPR derived from the Nellcor finger PPG and that from the Xhale finger PPG by shifting. Since PTT is on the scale of 100ms, to further minimize the impact of poor signal quality on PTT, beyond the applied SQI, we visually inspect the PPG signals to eliminate misclassified high-quality segments. ECG signal quality is also visually assessed, and R-peaks corresponding to RRI shorter than 0.25 s or longer than 1.5 s are excluded. Only time intervals with both high-quality PPG and ECG signals are retained for further PTT analysis. Following these preprocessing steps, the PTT of the *j*th cardiac cycle is calculated as the time difference between the R peak in the ECG signal and the **msp** of the associated PPG cycle. Since PTT tends to decrease with shorter RRIs and the baseline HR depends on subjects, we normalize the PTT to ensure comparability across individuals by their baseline RRI, that is, 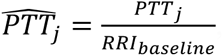, where *RRI_baseline_* is the median RRI during the baseline stage of the LBNP experiment.

For each of the above indices derived from each channel, including the SQI, we report the median and median absolute deviation (MAD) as it is more robust to outliers than the mean and standard deviation. For the demographic data, we report the mean and standard deviation (SD). We apply the Kruskal-Wallis test to test the hypothesis that the medians of an index derived from different body locations are equal against the alternative hypothesis that at least one body location gives a different result from the others. To compare two methods to define an index, we apply the Wilcoxon signed-rank test. We view a p-value less than 0.005 as statistically significant [30], and use the Bonferroni correction to handle the multiple test comparison. For example, since we compare 30 indices in total with Kruskal-Wallis test, the corrected p-value is 0.005/30=1.67×10^−4^.

## 4. Results

The dataset consists of 36 subjects (17 males and 19 females) subjects ages 18-40 underwent progressive LBNP protocol. Case #3 is missing. The overall signal length is 29.86±7m. Two flawed cases (one female “case #10” and one male “case #13”) were removed due to at least one broken channel. Thus, 33 cases were included in the analysis. The period of 30 m to 31.67 m of case #1, the period of 25 m to 27.42 m of case #7, the period of 29.5 m to the end of case #9, the first 4.5 m of case 25, the period of 29.3 m to 34.5 m of case #28, the period of 30 m to 31.67 m of case #29 and the first 3.8 m of case #32 are removed due to broken signals.

In the PTT analysis, we accounted for the quality of both Xhale and Necllor devices. Besides case #10 and case#13, cases #1, #5, #6, #7, #8, #18, #19, #25, and #27 were further excluded since we fail to align Nellcor and Xhale using **msp**-IPR and case #30 was excluded as the LBNP experiment was terminated early. In total, 23 cases are included in PTT analysis.

The overall SQI of PPG recorded from different body locations is 0.99±0.01 (0.98±0.02 and 0.98±0.01 respectively) for nasal (ear and finger respectively). The median and MAD of SQI for each case are reported in Table 2. The null hypothesis that the signal quality scores are the same for different body locations is rejected by Kruskal-Wallis test (p<10^−10^). Using 0.9 as the SQI threshold, 84.64% of all 5-second segments from nasal PPG signals were considered high quality, compared to 77.62% from ear and 81.35% from finger PPG signals across all subjects. The percentages of cases and sensor types with an SQI above this threshold are reported in Table 2. The progression of SQI over various LBNP experimental phases over different body locations is reported in Figure 4 with the *p*-value comparison reported in Table 3. We cannot reject that overall SQI is similar across different body locations at the −75 mmHg phase. All results described below assume only signal segments with an SQI of at least 0.9 are considered.

**Table 2:** Summary statistics of SQI for each subject. SQI is calculated after broken segments are removed. The second to fifth columns are the median±MAD of SQI, and the sixth to eighth columns are the ratios of high SQI (>0.9).

| Case | Nasal<br>(a.u) | Ear (a.u) | Finger<br>(a.u) | Nasal<br>(%) | Ear<br>(%) | Finger<br>(%) |
| --- | --- | --- | --- | --- | --- | --- |
| 1 | 0.99±0.00 | 0.99±0.01 | 0.99±0.01 | 87.63 | 87.37 | 90.79 |
| 2 | 0.98±0.01 | 0.93±0.05 | 0.97±0.01 | 88.63 | 61.50 | 89.41 |
| 4 | 0.99±0.01 | 0.98±0.01 | 0.99±0.01 | 80.93 | 84.47 | 73.02 |
| 5 | 0.98±0.01 | 0.96±0.03 | 0.97±0.02 | 74.87 | 69.54 | 70.30 |
| 6 | 0.99±0.00 | 0.99±0.00 | 0.99±0.00 | 98.95 | 95.03 | 96.60 |
| 7 | 0.99±0.00 | 1.00±0.00 | 0.77±0.05 | 89.70 | 87.80 | 3.25 |
| 8 | 0.98±0.01 | 0.91±0.06 | 0.99±0.00 | 82.44 | 53.32 | 97.64 |
| 9 | 0.98±0.01 | 0.98±0.02 | 0.98±0.01 | 91.81 | 80.23 | 89.55 |
| 11 | 0.99±0.01 | 0.99±0.01 | 0.99±0.00 | 78.72 | 79.01 | 95.63 |
| 12 | 0.99±0.00 | 0.99±0.00 | 0.99±0.00 | 93.88 | 95.72 | 97.55 |
| 14 | 0.99±0.00 | 0.99±0.01 | 0.99±0.00 | 96.86 | 90.00 | 99.43 |
| 15 | 0.95±0.02 | 0.92±0.05 | 0.92±0.03 | 84.51 | 57.04 | 60.92 |
| 16 | 0.99±0.01 | 0.80±0.10 | 0.98±0.01 | 94.70 | 16.51 | 94.08 |
| 17 | 1.00±0.00 | 1.00±0.00 | 0.80±0.17 | 92.18 | 95.24 | 34.69 |
| 18 | 0.97±0.02 | 0.97±0.02 | 0.87±0.08 | 78.06 | 79.31 | 38.24 |
| 19 | 0.99±0.00 | 0.98±0.01 | 0.69±0.19 | 95.74 | 82.98 | 20.57 |
| 20 | 0.99±0.01 | 0.98±0.01 | 0.98±0.01 | 94.79 | 82.99 | 95.14 |
| 21 | 0.99±0.00 | 0.93±0.06 | 0.99±0.01 | 94.99 | 57.23 | 89.09 |
| 22 | 0.99±0.00 | 0.96±0.03 | 0.98±0.01 | 98.41 | 71.75 | 85.71 |
| 23 | 0.99±0.01 | 0.98±0.01 | 0.98±0.02 | 79.63 | 91.90 | 72.69 |
| 24 | 0.99±0.00 | 0.97±0.02 | 0.99±0.00 | 97.16 | 80.85 | 99.29 |
| 25 | 0.99±0.01 | 0.84±0.09 | 0.99±0.00 | 96.31 | 28.11 | 100.00 |
| 26 | 0.99±0.00 | 0.97±0.02 | 0.99±0.00 | 94.08 | 74.67 | 99.67 |
| 27 | 0.93±0.06 | 0.95±0.05 | 0.79±0.14 | 54.95 | 60.75 | 28.33 |
| 28 | 0.37±0.26 | 0.99±0.01 | 0.98±0.01 | 21.04 | 96.30 | 92.89 |
| 29 | 0.99±0.00 | 0.99±0.01 | 0.99±0.01 | 87.66 | 87.14 | 90.81 |
| **30** | 0.99±0.01 | 0.73±0.03 | 0.98±0.01 | 88.61 | 0.00 | 95.02 |
| **31** | 0.99±0.01 | 1.00±0.00 | 0.99±0.00 | 91.27 | 99.44 | 92.39 |
| **32** | 0.99±0.00 | 0.99±0.00 | 0.99±0.01 | 96.34 | 93.29 | 99.09 |
| **33** | 1.00±0.00 | 1.00±0.00 | 0.99±0.00 | 97.02 | 96.03 | 98.34 |
| **34** | 0.98±0.01 | 0.99±0.01 | 0.98±0.01 | 88.55 | 89.16 | 94.58 |
| **35** | 0.99±0.00 | 1.00±0.00 | 0.99±0.00 | 92.44 | 98.04 | 91.60 |
| **36** | 0.98±0.01 | 0.98±0.01 | 0.99±0.01 | 84.08 | 87.15 | 85.47 |
| **All** | 0.99±0.01 | 0.98±0.02 | 0.98±0.01 | 84.64 | 77.62 | 81.35 |

**Table 3:** p-values (Kruskal-Wallis test) for SQI comparison over different experimental phases. * indicates we reject the null hypothesis that group medians are the same with statistical significance (p < 0.005/7=7.1×10^−4^ using the Bonferroni correction).

| **Experimental Stages** | Kruskal-Wallis test |
| --- | --- |
| **Baseline** | $<10^{-10} *$ |
| **-15** | $<10^{-10} *$ |
| **-30** | $<10^{-10} *$ |
| **-45** | $<10^{-10} *$ |
| **-60** | $6.01 \times 10^{-8} *$ |
| **-75** | $1.5 \times 10^{-3}$ |
| **Recovery** | $3.45 \times 10^{-7} *$ |

Next, we explored **dic**. The cardiac cycles with detectable **dic** are overall 43.68 ± 20.89% (40.20 ± 12.80% and 58.22 ± 8.04%) of all cardiac cycles in the nasal (ear and finger respectively) PPG. The ratios of cardiac cycles with detectable **dic** are different over different body locations with statistical (p =3.6×10^−3^). In terms of number of cardiac cycles, there are 28,549 (21,103 and 38,240 respectively) in the nasal (ear and finger respectively) PPG that have detectable **dic** and 37,937 (39,764 and 25,775 respectively) that are **dic**-free.

We then noted the relationship between the **dic** and e point over cardiac cycles with detectable **dic** from different body locations, which is plotted in Figure 5. For cardiac cycles where the **dic** is detectable, the root mean square of the time interval between the dic and the e-point is 20 ms for finger PPG, 15 ms for ear PPG, and 21 ms for nasal PPG; the corresponding medians are 14 ms, 6 ms, and 14 ms, respectively. In 97.86% (77.12% and 87.14% respectively) of the cardiac cycles with **dic**, the e point is strictly before the **dic** in the nasal (ear and finger respectively) PPG with the median time difference 16 ± 8 ms (10 ± 6 ms and 16 ± 8 ms respectively). In 1.22% (8.61% and 4.69% respectively) of these cycles, the e point is exactly at the **dic** in the nasal (ear and finger respectively) PPG, and in the remaining 0.93% (14.27% and 8.17% respectively), the e point is strictly after the **dic** in the nasal (ear and finger respectively) PPG with the median time difference 2 ± 0 ms (4 ± 0 ms and 4 ± 0 ms respectively). The time discrepancy among different channels is significantly different among different body locations when tested by the Kruskal-Wallis test (p < 10^−10^).

**Figure 5:**
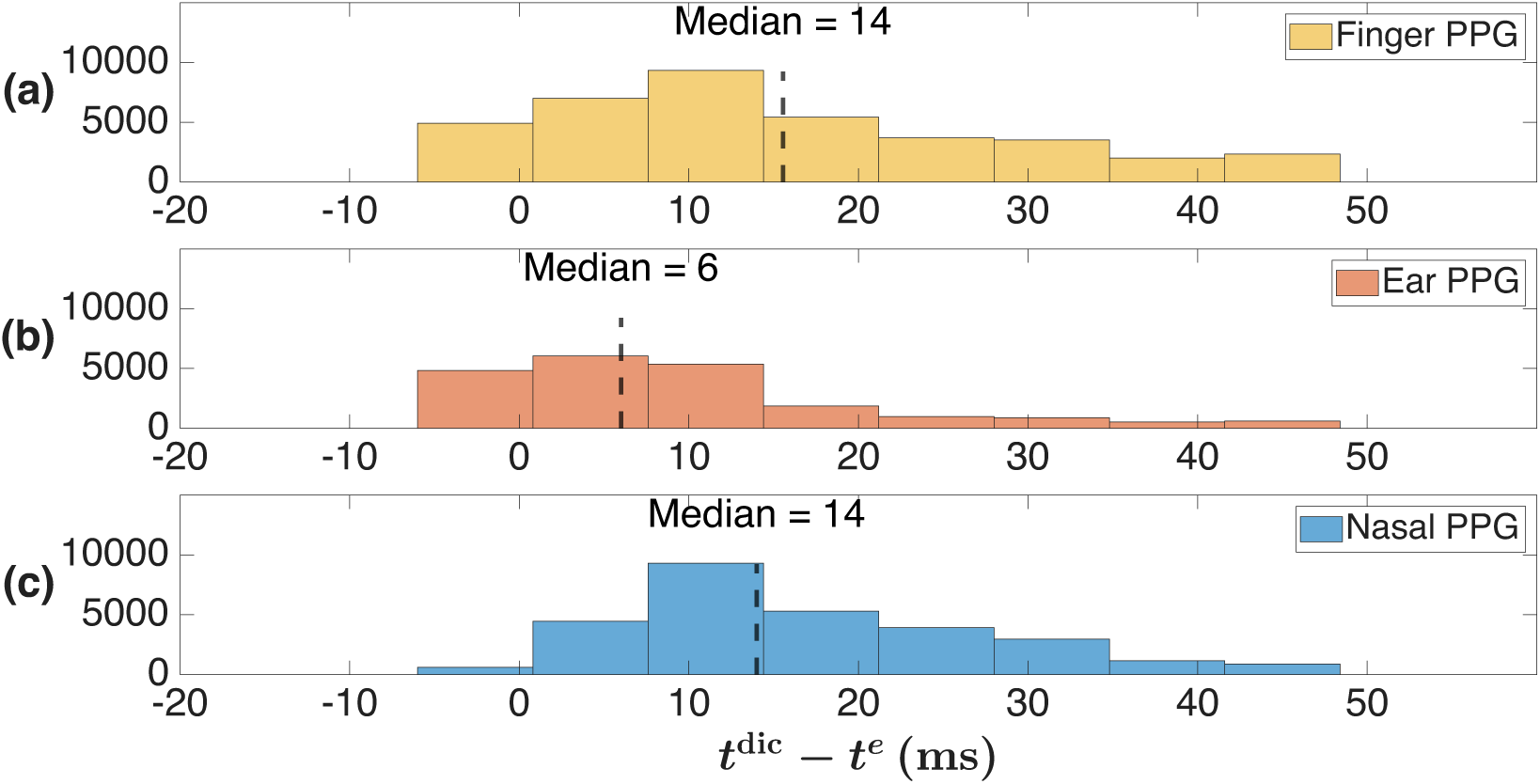
Histograms for the time difference between **dic** (*t*^dic^) and the e point (*t*^e^) for **(a)** Finger PPG, **(b)** ear PPG, and **(c)** nasal PPG.

We then explored the relationship between **dic** and other features. For those cardiac cycles with detectable **dic**, the time difference between **msp** and **dic** is 224 ± 36 ms (224 ± 42 ms and 262 ± 34 ms respectively) in the nasal (ear and finger respectively) PPG. These time differences were different over different body locations with statistical significance (p < 10^−10^). The time difference between **msp** and e point over all cycles is 208 ± 32 ms (218 ± 44 ms and 246 ± 34 ms respectively) in the nasal (ear and finger respectively) PPG. The time difference between **msp** and e in all cardiac cycles are different over different body locations with statistical significance (p < 10^−10^).

Continuing, we explored feature-related indices. The summary statistics of these indices derived from different body locations are shown in Table 4. In short, all indices are significantly different when evaluated from PPG signals recorded from different body locations, except PR. The **msp**-PR is 1.37 ± 0.17 bps (1.39 ± 0.18 bps and 1.39 ± 0.17 bps respectively) and the **sys**-PR is 1.37 ± 0.17 bps (1.39 ± 0.18 bps and 1.39 ± 0.17 bps respectively) from the nasal (ear and finger respectively) PPG. For both **msp**-PR and **sys**-PR, there is no statistically significant difference among body locations, as shown in Table 4. Using the Wilcoxon signed rank test comparing PR defined by **msp** and **sys** concludes that **msp**-PR and **sys**-PR were not significantly different in all body locations (p = 0.99, p = 0.86, and p = 0.99 in nasal, ear, and finger respectively).

**Table 4:**
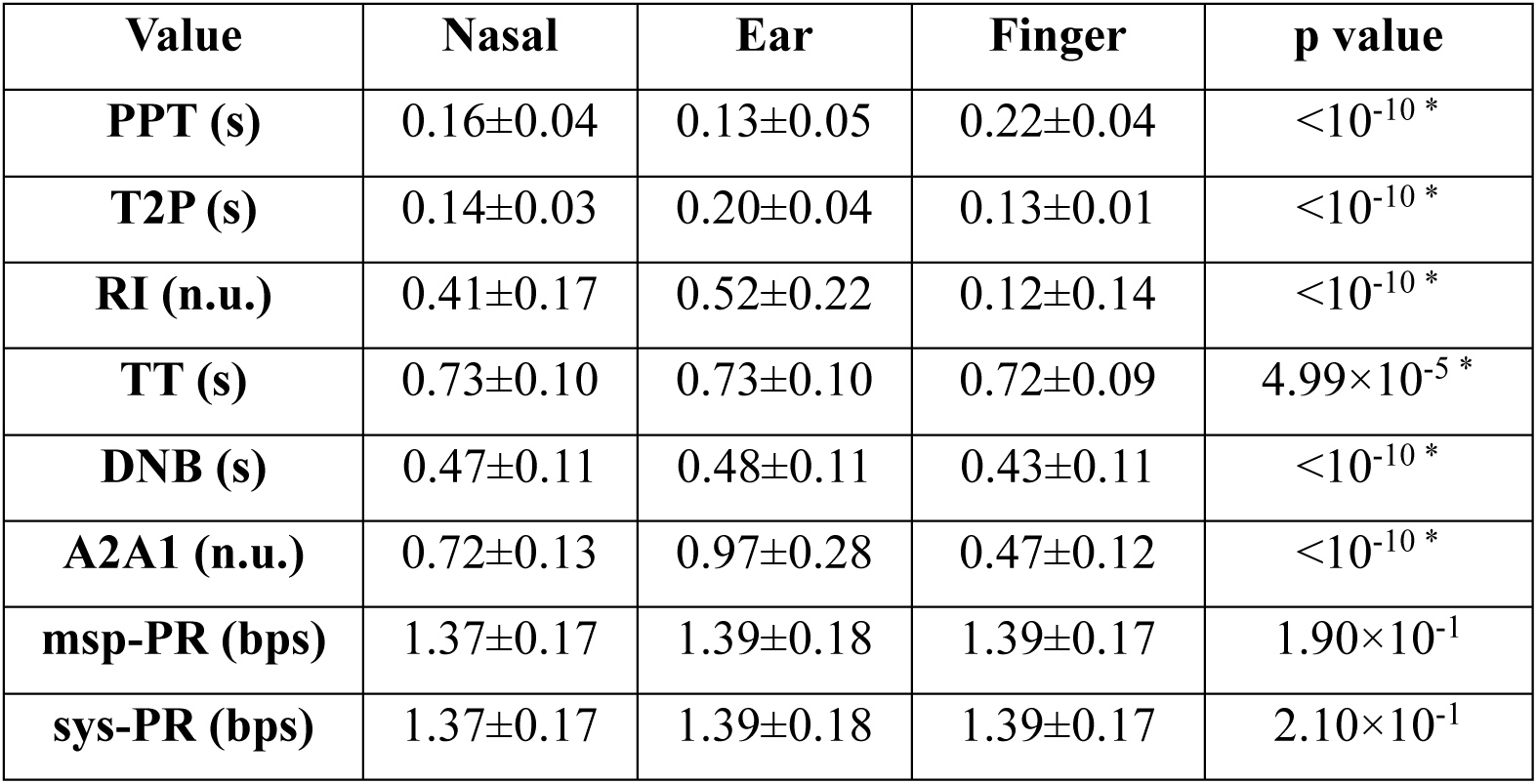
Summary statistics of the considered feature-related indices. All values are Median ± MAD and are taken for cycles with an SQI above or equal to 0.9. * indicates rejecting the null hypothesis that medians are equal (p<0.005/30=1.67×10^−4^).

| Value | Nasal | Ear | Finger | p value |
| --- | --- | --- | --- | --- |
| **PPT (s)** | 0.16±0.04 | 0.13±0.05 | 0.22±0.04 | $<10^{-10}$ * |
| **T2P (s)** | 0.14±0.03 | 0.20±0.04 | 0.13±0.01 | $<10^{-10}$ * |
| **RI (n.u.)** | 0.41±0.17 | 0.52±0.22 | 0.12±0.14 | $<10^{-10}$ * |
| **TT (s)** | 0.73±0.10 | 0.73±0.10 | 0.72±0.09 | $4.99 \times 10^{-5}$ * |
| **DNB (s)** | 0.47±0.11 | 0.48±0.11 | 0.43±0.11 | $<10^{-10}$ * |
| **A2A1 (n.u.)** | 0.72±0.13 | 0.97±0.28 | 0.47±0.12 | $<10^{-10}$ * |
| **mSP-PR (bps)** | 1.37±0.17 | 1.39±0.18 | 1.39±0.17 | $1.90 \times 10^{-1}$ |
| **sys-PR (bps)** | 1.37±0.17 | 1.39±0.18 | 1.39±0.17 | $2.10 \times 10^{-1}$ |

Next, we report the exploration of PRV related indices. The **msp**-SDNN was 74 ± 21 ms (80 ± 26 ms and 65 ± 18 ms respectively) and the **sys**-SDNN was 74 ± 21 ms (85 ± 26 ms and 66 ± 19 ms respectively) for the nasal (ear and finger respectively) PPG. Using the Wilcoxon signed rank test comparing SDNN defined by **msp** and **sys** concludes that **msp**-SDNN and **sys**-SDNN were not significantly different for nasal, ear, and finger PPGs (p = 0.87, p = 0.04 and p = 0.90 respectively). The **msp**-RMSSD was 63 ± 28 ms (76 ± 36 ms and 39 ± 17 ms respectively) and the **sys**-RMSSD was 63 ± 26 ms (88 ± 38 ms and 40 ± 18 ms respectively) for the nasal (ear and finger respectively) PPG. Using the Wilcoxon signed rank test comparing RMSSD defined by **msp** and **sys** concludes that **msp**-RMSSD and **sys**-RMSSD were not significantly different for nasal and finger PPG (p = 3.36×10^−2^, p = 6.88×10^−1^, respectively), while a significant difference was found for ear PPG (p = 8.16×10⁻⁵). Summary statistics for PRV related indices are reported in Table 5.

**Table 5:** Pulse rate variabilities evaluated using different features (**msp** and **sys**) derived from different body locations (nasal, ear and finger). All values are shown in Median ± MAD and are taken for cycles with an SQI above or equal to 0.9. The Wilcoxon rank test is used to compare the use of **msp** vs **sys** as indices under the null hypothesis of equal medians, where ^#^ indicates rejecting the null hypothesis that medians are equal (p<0.005/9=5.56×10^−4^). * indicates rejecting the null hypothesis that medians are equal by the Kruskal-Wallis test (p<0.005/30=1.67×10^−4^). n.u. means no unit.

|  | Nasal | Ear | Finger | p value |
| --- | --- | --- | --- | --- |
| **mSP-SDNN (ms)** | 74±21 | 80±26 | 65±18 | $7.58 \times 10^{-10}$ * |
| **sys-SDNN (ms)** | 74±21 | 85±26 | 66±19 | $<10^{-10}$ * |
| **p value (mSP vs sys)** | $8.71 \times 10^{-1}$ | $4.48 \times 10^{-2}$ | $9.02 \times 10^{-1}$ | |
|  | Nasal | Ear | Finger | p value |
| **mSP-RMSSD (ms)** | 63±28 | 76±36 | 39±17 | $<10^{-10}$ * |
| **sys-RMSSD (ms)** | 63±26 | 88±38 | 40±18 | $<10^{-10}$ * |
| **p-value (mSP vs sys)** | $3.36 \times 10^{-2}$ | $8.16 \times 10^{-5}$ # | $6.88 \times 10^{-1}$ | |
|  | Nasal | Ear | Finger | p value |
| **mSP-SD1 (ms)** | 44±19 | 54±26 | 27±12 | <10<sup>-10</sup> * |
| **sys-SD1 (ms)** | 45±19 | 62±27 | 28±13 | <10<sup>-10</sup> * |
| **p value (msp vs sys)** | 3.36×10<sup>-2</sup> | 8.24×10<sup>-5</sup> # | 6.87×10<sup>-1</sup> |  |
|  | **Nasal** | **Ear** | **Finger** | **p value** |
| **mSP-SD2 (ms)** | 92±25 | 99±29 | 85±23 | 3.06×10<sup>-4</sup> |
| **sys- SD2 (ms)** | 91±25 | 100±28 | 86±23 | 2.25×10<sup>-5</sup> * |
| **p value (msp vs sys)** | 6.53×10<sup>-1</sup> | 4.40×10<sup>-1</sup> | 9.38×10<sup>-1</sup> |  |
|  | **Nasal** | **Ear** | **Finger** | **p value** |
| **mSP-pNN50 (%)** | 6.24±4.64% | 10.34±8.39% | 5.65±4.77% | <10<sup>-10</sup> * |
| **sys- pNN50 (%)** | 12.07±7.83% | 26.28±16.55% | 6.26±5.36% | <10<sup>-10</sup> * |
| **p value (msp vs sys)** | <10<sup>-10</sup> # | <10<sup>-10</sup> # | 4.33×10<sup>-1</sup> |  |
|  | **Nasal** | **Ear** | **Finger** | **p value** |
| **mSP-pNN20 (%)** | 34.00±16.42% | 42.64±15.19% | 37.08±16.61% | <10<sup>-10</sup> * |
| **sys-pNN20 (%)** | 38.86±14.93% | 60.17±13.73% | 36.48±17.38% | <10<sup>-10</sup> * |
| **p value** | 2.53×10<sup>-4</sup> # | <10<sup>-10</sup> # | 7.51×10<sup>-1</sup> |  |
|  | **Nasal** | **Ear** | **Finger** | **p value** |
| **mSP-LF (ms<sup>2</sup>)** | 1263.80±691.45 | 1228.65±758.86 | 791.31±414.25 | <10<sup>-10</sup> * |
| **sys-LF (ms<sup>2</sup>)** | 1129.20±637.27 | 1219.26±739.96 | 786.69±414.88 | <10<sup>-10</sup> * |
| **p value (msp vs sys)** | 4.76×10<sup>-2</sup> | 6.97×10<sup>-1</sup> | 7.77×10<sup>-1</sup> |  |
|  | **Nasal** | **Ear** | **Finger** | **p value** |
| **mSP-HF (ms<sup>2</sup>)** | 1146.48±832.01 | 1521.34±1158.66 | 618.84±400.67 | <10<sup>-10</sup> * |
| **sys-HF (ms<sup>2</sup>)** | 1073.99±773.05 | 1755.87±1326.89 | 604.38±403.96 | <10<sup>-10</sup> * |
| **p value (msp vs sys)** | 2.88×10<sup>-1</sup> | 2.00×10<sup>-1</sup> | 4.85×10<sup>-1</sup> |  |
|  | **Nasal** | **Ear** | **Finger** | **p value** |
| **mSP-LHR (n.u.)** | 1.05±0.38 | 0.81±0.26 | 1.30±0.62 | <10<sup>-10</sup> * |
| **sys-LHR (n.u.)** | 1.03±0.35 | 0.74±0.25 | 1.33±0.63 | <10<sup>-10</sup> * |
| **p value (msp vs sys)** | 8.40×10<sup>-1</sup> | 3.64×10<sup>-3</sup> | 4.23×10<sup>-1</sup> |  |

We then evaluate harmonic related indices (PHI and AHI) and their statistics. See Table 6 for a summary. It shows that PHI and AHI evaluated from PPGs recorded from different body sites have significant difference [8, 21–24], which is consistent with the previous findings showing different oscillatory morphologies among PPGs recorded from different body locations.

**Table 6:** AHI and PHI evaluated derived from different body locations (nasal, ear and finger). All values are shown in Median ± MAD and are taken for cycles with an SQI above or equal to 0.9. * indicate rejecting the null hypothesis that medians are equal (p<0.005/30=1.67×10^−4^).

|  | **Nasal** | **Ear** | **Finger** | **p value** |
| --- | --- | --- | --- | --- |
| **PHI2 (radian)** | 1.77±0.72 | 2.16±0.95 | 1.36±0.71 | $< 10^{-10}$ * |
| **PHI3 (radian)** | 1.80±2.95 | -2.07±1.65 | 2.22±1.43 | $< 10^{-10}$ * |
| **AHI2 (a.u)** | 0.50±0.21 | 0.38±0.2 | 0.59±0.2 | $< 10^{-10}$ * |
| **AHI3 (a.u)** | 0.26±0.08 | 0.19±0.08 | 0.25±0.09 | $< 10^{-10}$ * |

Finally, we present the PTT analysis. The normalized PTT, calculated using Nellcor finger PPG, Xhale finger PPG, Xhale ear PPG, and Xhale nose PPG, is 0.30±0.02, 0.30±0.03, 0.23±0.03, and 0.21±0.03 (no unit), respectively. First, we show the PTT derived from Nellcor finger PPG and ECG across different LBNP stages for all subjects in the first panel of Figure 7. We cannot see a clear relationship between normalized PTT and LBNP phases. Second, we explore the PTT derived from different body locations in the second to fourth panels in Figure 7. We find statistically significant difference between the results using Nellcor finger PPG and Xhale finger PPG (p<10^−10^), while the absolute error between these two PTTs is 4ms±2ms. The normalized PTT derived from Xhale finger, ear and nose PPGs are statistically different (p<10^−10^). Figure 8 shows the unnormalized PTT determined from Nellcor finger, Xhale finger, Xhale ear, and Xhale nose PPGs, which are statistically different (p<10^−10^).

**Figure 6:**
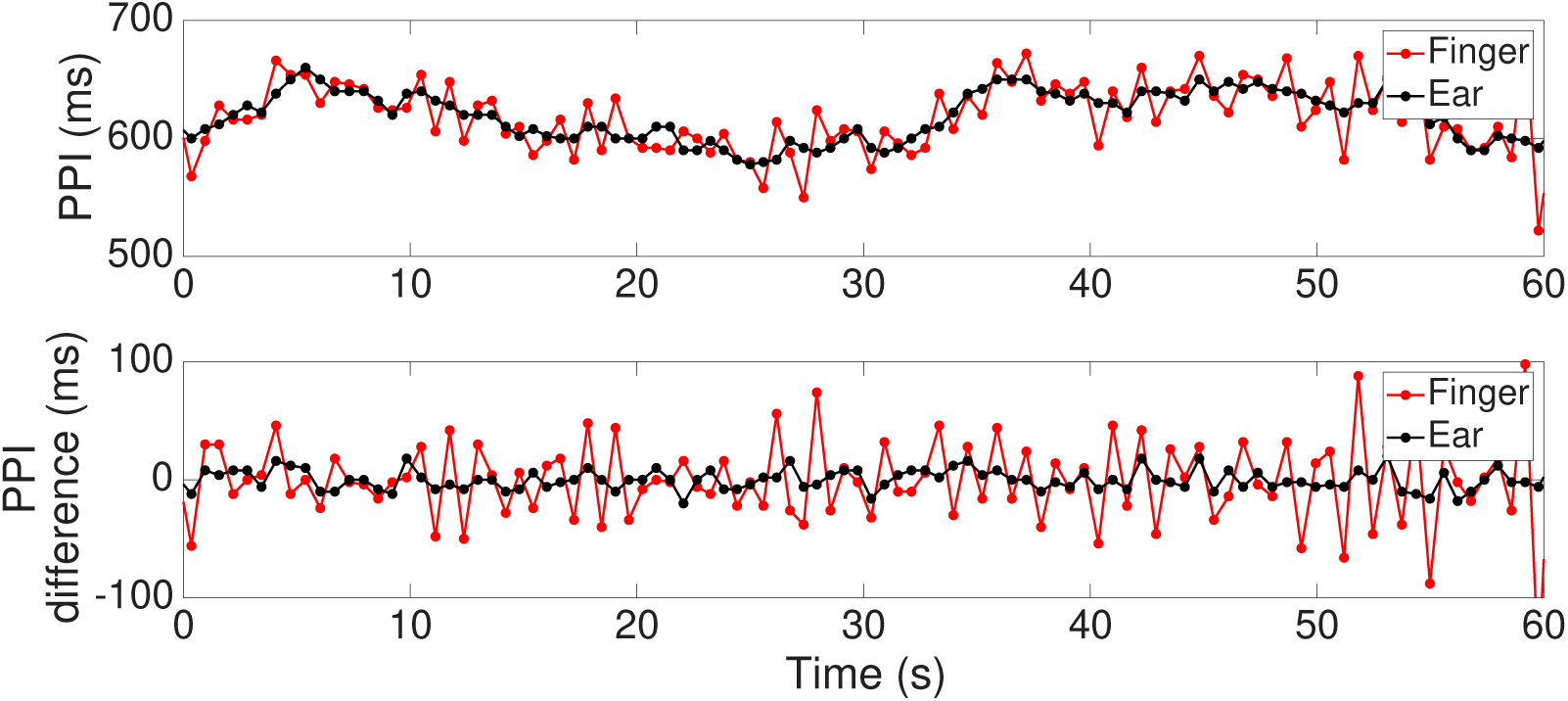
An illustration of pulse-to-pulse interval (PPI) shown in (a) and its derivative (dPPG) shown in (b). The red (black respectively) curves are PPI and dPPI derived from the finger (ear respectively) PPG.

**Figure 7:**
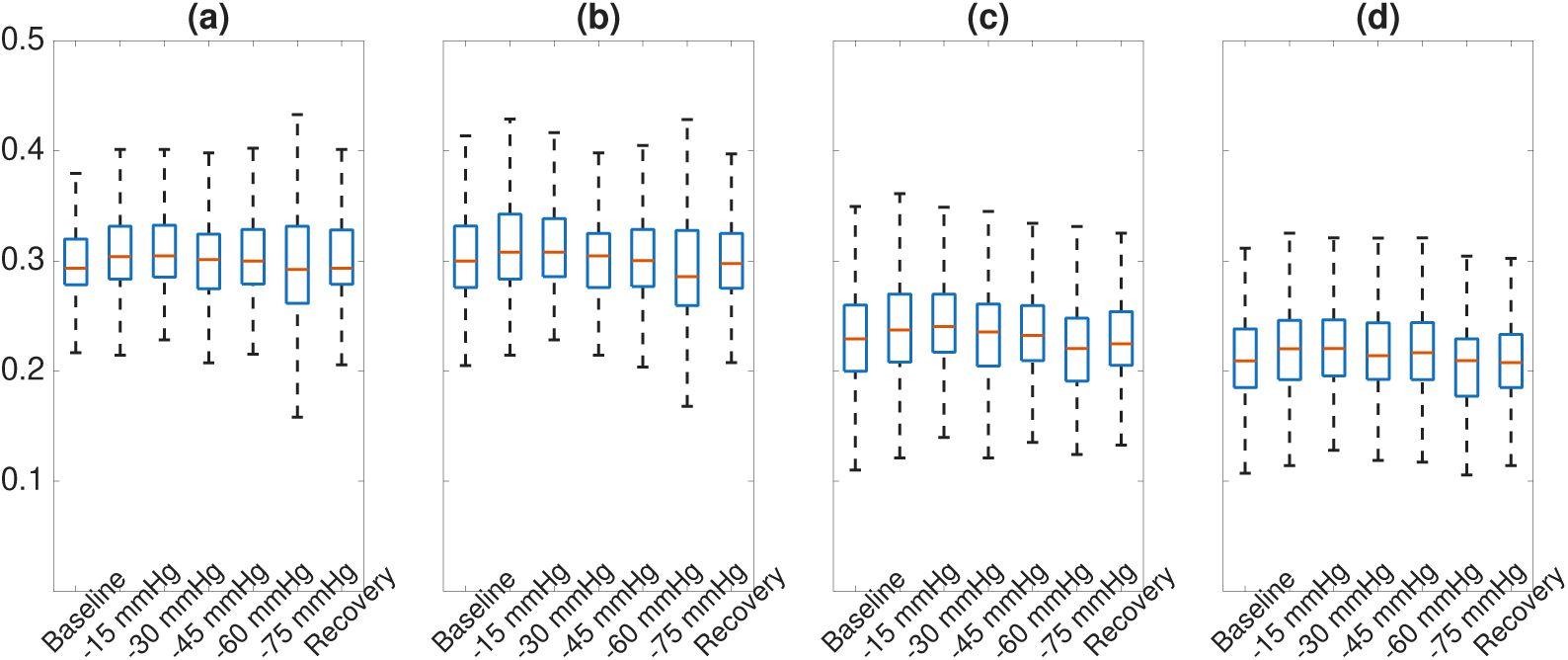
The boxplot of normalized PTT across different locations and LBNP stages. From left to right: Boxplots of normalized PTT across different LBNP stages, measured using(a)Nellcor finger PPG, (b)Xhale finger PPG, (c)Xhale ear PPG and (d)Xhale nasal PPG. The normalized PTT, calculated using Nellcor finger PPG, Xhale finger PPG, Xhale ear PPG, and Xhale nose PPG, is 0.30±0.02, 0.30±0.03, 0.23±0.03, and 0.21±0.03 (no unit), respectively.

**Figure 8:**
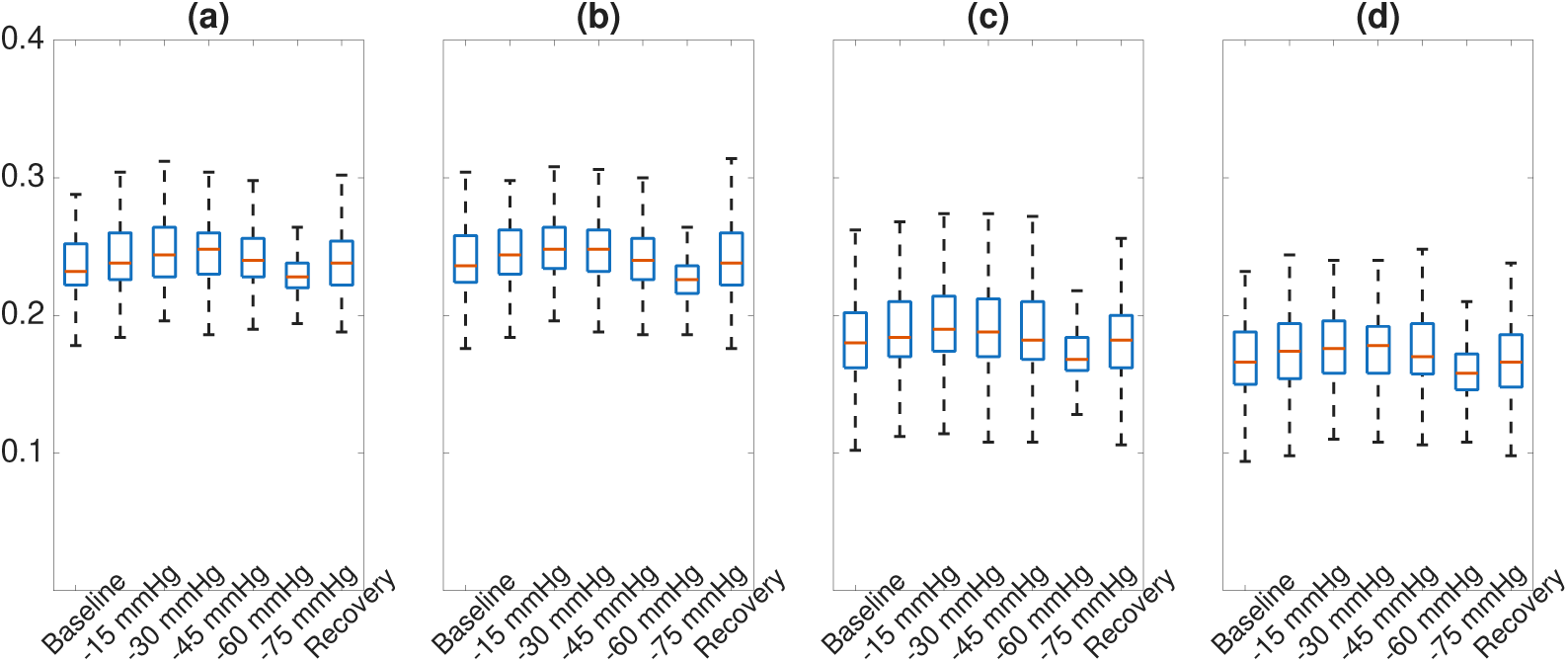
The boxplot of non-normalized PTT across different locations and LBNP stages. From left to right: Boxplots of non-normalized PTT across different LBNP stages, measured using (a) Nellcor finger PPG, (b) Xhale finger PPG, (c) Xhale ear PPG and (d) Xhale nasal PPG, and Nellcor EKG. The non-normalized PTT, calculated using Nellcor finger PPG, Xhale finger PPG, Xhale ear PPG, and Xhale nose PPG, is 0.24±0.01, 0.24±0.02, 0.18±0.02, and 0.17±0.02 (sec), respectively.

## 5. Discussion and conclusion

In this paper, we study a fundamental question of whether indices derived from PPG signals recorded from different body locations are equivalent? We have several findings. First, signal quality is not perfect even when PPG recordings are collected under well-controlled experimental conditions, and it varies across different measurement sites. Among the non-broken segments of all recordings listed in Table 2, the median and MAD of the high-SQI ratios across subjects are 91.27% ± 5.04% for nasal, 82.99% ± 11.24% for ear, and 91.60% ± 6.04% for finger PPG. These results suggest that finger PPG provides the highest signal quality, with nasal PPG showing comparable performance, while ear PPG yields the lowest quality. However, the Kruskal-Wallis test indicates that these differences are not statistically significant (p=0.051). This pattern might be explained by physiological and mechanical factors. The nasal region is richly perfused and less susceptible to motion or peripheral vasoconstriction, which contributes to stable signal quality. Similarly, the finger often provides strong PPG signals due to good vascular supply [47], though it can be more vulnerable to motion artifacts or circulation variability in some conditions. In contrast, the ear is generally more prone to such disturbances, particularly under low-perfusion states. Although statistical testing did not show a significant difference, the observed trend underscores the sensitivity of PPG signal quality to sensor placement, which is an important consideration in both clinical monitoring and the development of wearable health technologies. In the online supplementary, we prepare a parallel set of tables when SQI>0.9 is not imposed for a comparison.

Second, most feature-related indices and PHI and AHI are different among body locations with statistical significance, except pulse rate (PR). Note that PPGs recorded from all body locations have different ratios of detectable dicrotic notch (**dic**), with finger PPG being the highest and ear PPG as the lowest. When **dic** is detectable, the e point and **dic** are also significantly different between different locations.

Since **dic** has a critical role in many applications, for example, in **dic** to baseline time (DNB), reflective index (RI) [13, 14] and stiffness index (SI) [15, 16], it is worth further discussion. There have been several algorithms proposed to automatically identify **dic**. To our knowledge, one of the earliest relevant algorithms aiming to detect **dic** in the brachial arterial pressure waveform was proposed in 1968 [31]. Several algorithms followed, including a compression scheme [32], a successive chord approach [33], the second derivatives of the waveform approach [34, 35], a rule-based logic approach [11], a nonconvex optimization approach [36, 37], a model based approach [20], physics-aware approach [52], etc. In general, there is no guarantee that a well-defined **dic** exists in a cardiac cycle, and when such a notch is not detectable, a dilemma appears. That is, since **dic** is *originally* defined in the time-domain by the notch by a specific morphological description, if such notch does not exist, what is **dic**? This often depends on the researcher’s discretion. A traditional approach when the **dic** is undetectable is using the e point determined from the APPG as a surrogate of **dic**. However, the exploration in this paper suggests that using the e point might not be the optimal surrogate when **dic** does not exist. Mathematically, this is because the e point is the locally fastest changing point of the first derivative of PPG, while **dic** is the 0 point of the first derivative of PPG, and they may sit in different locations. At first glance, the roughly 15∼20 millisecond difference between the e point and **dic** when **dic** exists might be harmless in clinical applications, and it is always possible to replace **dic** by the e point systematically to avoid this discrepancy. However, it does not resolve the “what is **dic**” problem. A recent effort using a decomposition scheme [38] to determine the **dic** might alleviate this issue. From mathematical perspective, this decomposition could be modeled by the adaptive non-harmonic model framework [8] so that the fundamental component is suppressed and the **dic** is enhanced. While this is potential in resolving the issue of replacing **dic** by the e point, the dependence of **dic** on PPG sensor locations, and the understanding of its mechanism, still exists. We need more work to handle this challenging problem, and we will discuss it to our future work.

The study results coincide with the well-known fact that PPG recorded from different body locations capture different physiological dynamics, which is represented in the PPG morphology [8, 9, 21, 24]. For example, we show that there are significantly more cardiac cycles (and higher ratio) in the finger PPG that have detectable **dic** compared with the ear and nasal PPGs. The time difference between **msp** and **dic** for those cardiac cycles with detectable **dic** is significantly different in these three PPG signals. Moreover, we shall mention that PHI and AHI are evaluated using the time-frequency analysis of PPG, which encodes PPG dynamics jointly from the time and frequency domains. From the anatomical perspective, the ear, nose, and finger differ in arterial supply, vascular density, and proximity to core circulation [50], all of which can influence PPG signal morphology and derived indices. For example, the finger PPG is more influenced by peripheral vasoconstriction and thermal regulation due to its distal location and high surface-area-to-volume ratio, while the nasal and ear sites, supplied by branches of the external and internal carotid arteries, may be closer representations of central circulation under resting conditions. Additionally, regional differences in arterial compliance, microvascular structure, and local sympathetic innervation [51] can affect waveform characteristics such as the dicrotic notch, amplitude, and timing features. These anatomical and hemodynamic differences offer a plausible basis for the site-specific variation observed in our analysis. These findings, combined with existing literature, further confirm that PPG recorded from different body locations are different and should not be viewed as the same signal. We hypothesize that by integrating PPGs recorded from different body locations, we may obtain a more systematic understanding of the hemodynamics. We will explore this possibility in our future study.

Since PRs evaluated from PPGs recorded from different body locations are not statistically different, the discrepancy of PRV with statistical significance may initially seem concerning and deserve a discussion. Particularly, PRV indices evaluated from finger PPG are smaller than those evaluated from nasal or ear PPG. From a physiological perspective, the finger has a strong blood perfusion and hence a more consistent vascular tone [49, 50]. Additionally, finger PPG is typically more mechanically stable compared to ear or nasal PPG. Both might explain the findings. This discrepancy also involves PTT. A recent study [42] reports that for individuals with greater PTT variability, PRV is not a good surrogate for HRV. From a mathematical perspective, note that if we view a sequence of pulse-to-pulse intervals (PPI) as a random process, the PR is its mean, and SDNN is its standard deviation. These two quantities are of different nature, so a similar PR does not imply a similar SDNN. RMSSD is the root mean square of successive PPI difference, which has a different nature compared with PPI since the finite difference operator is applied. Moreover, it is important to notice that the difference operator could amplify the discrepancy among PPIs evaluated from different PPGs. See Figure 6 for an example, where PPIs from finger-PPG and ear-PPG and their differences, denoted as dPPI, are shown. The mean and SD of the difference of these two PPIs are <10^−4^ ms and 1.177 ms, and the mean and SD of the difference of these two dPPIs are −0.004 ms and 2.078 ms. As a result, it is not surprising that PRV-related indices are significantly different when evaluated from different PPIs. Given that PRV-related indices serve as crucial alternatives to HRV indices when electrocardiogram data is unavailable [39], researchers should be careful when interpreting these indices. This caution is particularly important when extrapolating results obtained from PRV-related indices evaluated from one body’s location to those evaluated from a different body location.

In this study, the use of the nose as a measurement site was also included, although it is not a standard location for transmissive PPG. This approach represents a novel application of PPG sensors, initially introduced by Xhale Inc. It is particularly useful for obtaining reliable waveforms in patients with compromised peripheral circulation, such as those with Raynaud’s phenomenon or those receiving high doses of vasopressors.

The findings related to PTT require further discussion. In this study, PTT measured at the ear and nose are significantly shorter than that measured at the finger, likely due to anatomical differences; that is, the arterial path from heart to the finger is longer than to the ear or nose. Notably, we observe a statistically significant difference between PTT measured using the Nellcor finger PPG and that from the Xhale finger PPG, despite synchronizing the devices via IPR alignment. This highlights a key limitation of our PTT analysis: ECG and PPG signals were recorded using separate devices without a shared hardware clock for synchronization. Although IPR was used for temporal alignment, discrepancies between hardware systems may still introduce measurement errors. Furthermore, normalized PTT exhibited limited sensitivity to LBNP phases, even when using ECG and PPG signals both acquired from the Nellcor device. This result contrasts with previously published findings. Therefore, while we can confirm that PTT varies with measurement site, we are unable to draw stronger conclusions. It is also important to note that the Nellcor system operates as a black box [40], and we cannot rule out the possibility that its ECG and PPG signals are not synchronized sufficiently for accurate PTT analysis, as has been reported for other commercial systems [41].

In this study, all PPGs are recorded when the subjects are resting during a standardized LBNP experiment, which is a well-controlled condition. A natural question that arises is whether the same conclusions hold for signals recorded from the clinical or free-living environments or reflective type PPG sensors that are increasingly common in real-world applications. From this perspective, the scope of the study is limited. However, the relationships established under controlled conditions serve as a necessary foundation for understanding how these indices behave in clinical or free-living environments, where not only is the SQI typically lower, but location-dependent motion artifacts and other outliers are also present. These factors may lead to erroneous indices that adversely affect the decision-making process. Furthermore, our analysis focuses on high-SQI signal segments. At first glance, this may seem restrictive. However, it is important to note that when SQI is low, the identification of physiological landmarks becomes unreliable, resulting in indices and comparisons that lack meaningful interpretation. This reflects common practice, in which low-SQI segments are typically excluded from analysis to ensure data quality and reliability.

There are more limitations in this work. We only explore part of the available indices researchers use in practice, and the conclusion may not be extrapolated to the unexplored indices. The population is limited to young subjects. Are the same conclusions carried over to older subjects? We focus on contact PPG placed in finger, nasal and ear. The conclusion cannot be extrapolated to other contact PPG locations; for example, toe [43], forehead or foot [44], nor noncontact PPG [45]. Moreover, while it is intuitively correct that features depend on body locations with anatomical explanation, the physiological mechanisms are not yet fully known. Exploring these interesting topics will be reported in our future work.

## 6. Conclusion

In this study, we show that while certain indices derived from PPG signals, for example, pulse rate (PR) and the reflective index (RI), remain relatively stable across different body locations, many others are significantly influenced by sensor placement, even under well-controlled experimental conditions with minimal noise or artifacts. For instance, the e-point, commonly used as a surrogate marker for the dicrotic notch, may not generalize across body sites. Although our analysis does not cover all possible indices, and the findings may not be directly applicable to typical clinical settings or reflective-type PPG sensors, they clearly highlight the sensitivity of signal features to measurement location. We propose that PPG signals recorded from different body sites be regarded as highly correlated, yet distinct, channels and conclude that indices derived from PPG depend on where the PPG sensor is placed in the body. Researchers should be careful when applying conclusions established from PPG recorded from one body location to another PPG recorded from a different body location.

## Data Availability

All data produced in the present study are available upon reasonable request to the authors

## Online supplementary

Comparison between features from PPG with or without SQI selection

A. Dicrotic notch

1. Percentage of cardiac cycles with detectable dicrotic notch (nasal, ear, finger)

i. with SQI>0.9: 43.68 ± 20.89% (40.20 ± 12.80% and 58.22 ± 8.04%)
ii. without SQI: 43.33 ± 23.50% (39.57 ± 14.86% and 57.51 ± 7.31%)
2. Number of cardiac cycles with detectable dicrotic notch vs dicrotic notch free.

i. with SQI>0.9:

1. detectable dic: 28,549 (21,103 and 38,240 respectively)
2. dic -free: 37,937 (39,764 and 25,775 respectively)
ii. without SQI:

1. detectable dic: 33,099 (29,914 and 48,546, respectively)
2. dic -free: 46,111 (48,080 and 31,390, respectively)
3. Time between e-point and dicrotic notch

i. RMSE (ms)

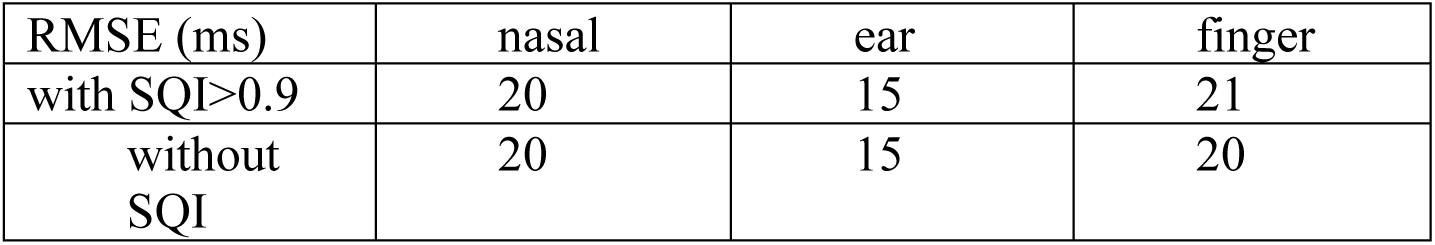
i. Percentage of cardiac cycles with e point strictly before the dicrotic notch, median time difference

| RMSE (ms) | nasal | ear | finger |
| --- | --- | --- | --- |
| with $SQI > 0.9$ | 20 | 15 | 21 |
| without $SQI$ | 20 | 15 | 20 |

- i. Percentage of cardiac cycles with e point strictly before the dicrotic notch, median time difference
| Percentage (%) | nasal | ear | finger |
| --- | --- | --- | --- |
| with $SQI > 0.9$ | 97.86 | 77.12 | 87.14 |
| without $SQI$ | 97.15 | 75.14 | 84.86 |

ii. Percentage of cardiac cycles with e point exactly at the dicrotic notch

| time difference (ms) | nasal | ear | finger |
| --- | --- | --- | --- |
| with $SQI > 0.9$ | $16 \pm 8$ | $10 \pm 6$ | $16 \pm 8$ |
| without $SQI$ | $16 \pm 8$ | $10 \pm 6$ | $14 \pm 8$ |

iii. Percentage of cardiac cycles with e point strictly after the dicrotic notch, median time difference

ii. Percentage of cardiac cycles with e point exactly at the dicrotic notch
| Percentage (%) | nasal | ear | finger |
| --- | --- | --- | --- |
| with SQI>0.9 | 1.22 | 8.61 | 4.69 |
| without SQI | 1.44 | 9.06 | 5.42 |

iii. Percentage of cardiac cycles with e point strictly after the dicrotic notch, median time difference
| Percentage (%) | nasal | ear | finger |
| --- | --- | --- | --- |
| with SQI>0.9 | 0.93 | 14.27 | 8.17 |
| without SQI | 1.41 | 15.80 | 9.71 |

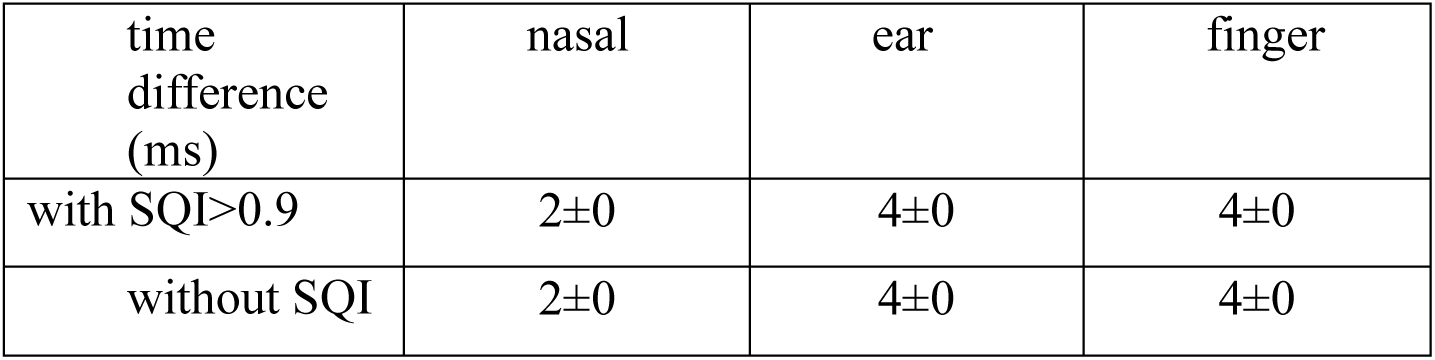

| time difference (ms) | nasal | ear | finger |
| --- | --- | --- | --- |
| with SQI>0.9 | 2±0 | 4±0 | 4±0 |
| without SQI | 2±0 | 4±0 | 4±0 |

B. MSP(Maximum slope point)

1. Time between MSP and dicrotic notch (ms)

2. Time between MSP and e-points (ms)

B. MSP(Maximum slope point) 1. Time between MSP and dicrotic notch (ms)
|  | nasal | ear | finger |
| --- | --- | --- | --- |
| with SQI>0.9 | 224± 36 | 224± 42 | 262± 34 |
| without SQI | 220 ±36 | 224 ±46 | 254± 40 |

2. Time between MSP and e-points (ms)
|  | nasal | ear | finger |
| --- | --- | --- | --- |
| with SQI>0.9 | 208± 32 | 218± 44 | 246 ±34 |
| without SQI | 206± 32 | 218± 46 | 240 ±38 |

C. Table 4 with and without SQI

1. Table 4 with SQI>0.9

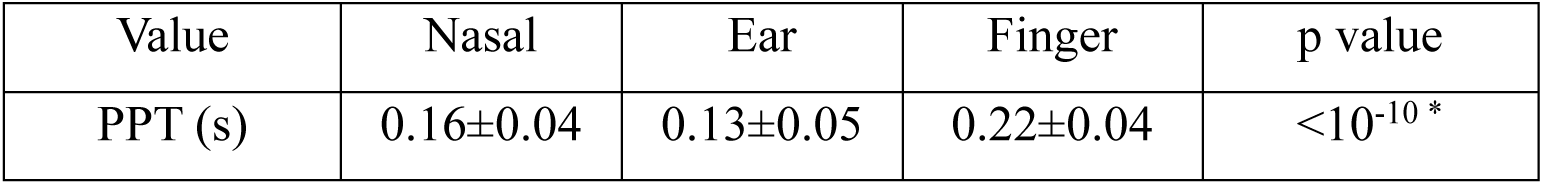
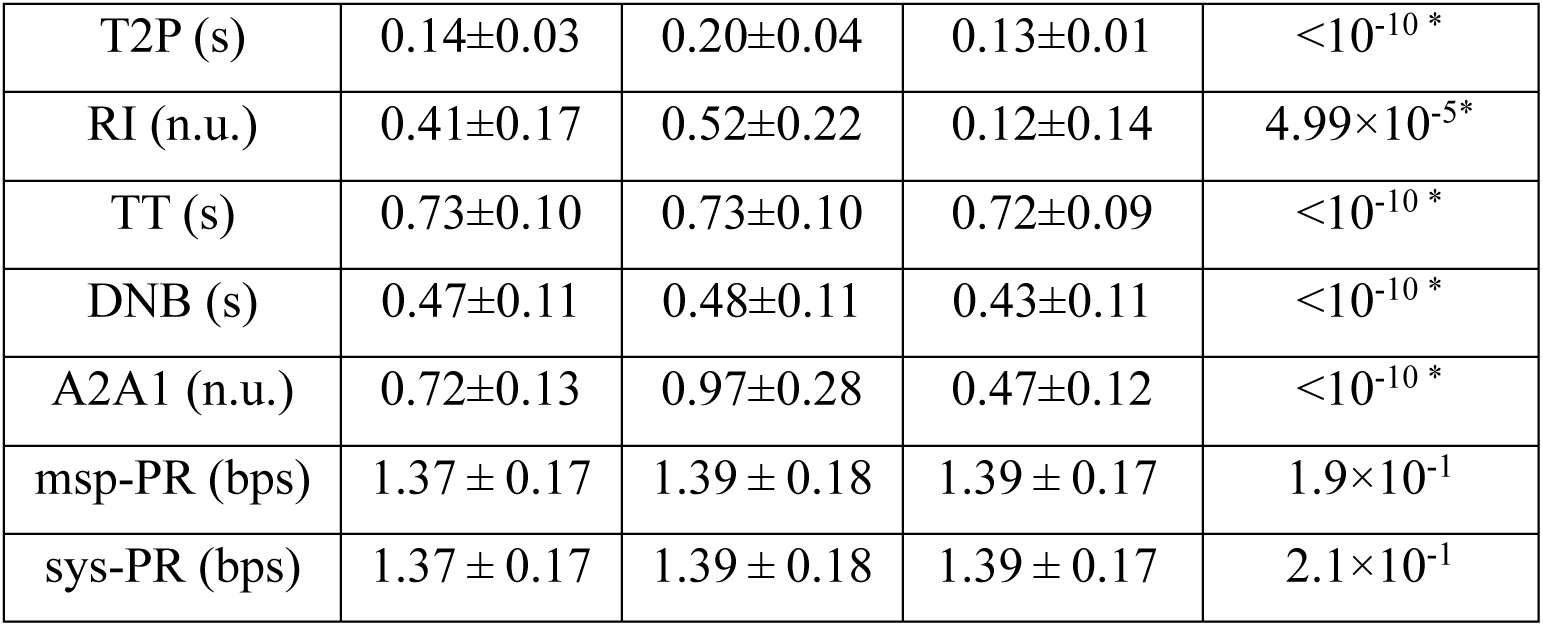
2. Table 4 without SQI

| Value | Nasal | Ear | Finger | p value |
| --- | --- | --- | --- | --- |
| PPT (s) | 0.16±0.04 | 0.14±0.06 | 0.22±0.05 | <10<sup>-10</sup> * |
| T2P (s) | 0.14±0.03 | 0.20±0.04 | 0.13±0.02 | <10<sup>-10</sup> * |
| RI (n.u.) | 0.39±0.17 | 0.51±0.25 | 0.12±0.15 | <10<sup>-10</sup> * |
| TT (s) | 0.71±0.10 | 0.72±0.10 | 0.71±0.10 | <10<sup>-10</sup> * |
| DNB (s) | 0.47±0.12 | 0.47±0.12 | 0.43±0.12 | <10<sup>-10</sup> * |
| A2A1 (n.u.) | 0.71±0.13 | 0.94±0.30 | 0.47±0.12 | <10<sup>-10</sup> * |
| msp-PR (bps) | 1.40±0.18 | 1.40±0.18 | 1.42±0.19 | 5.5×10<sup>-2</sup> |
| sys-PR (bps) | 1.40±0.18 | 1.40±0.18 | 1.42±0.19 | 4.8×10<sup>-2</sup> |

D. Table 5 with and without SQI

1. Table 5 with SQI

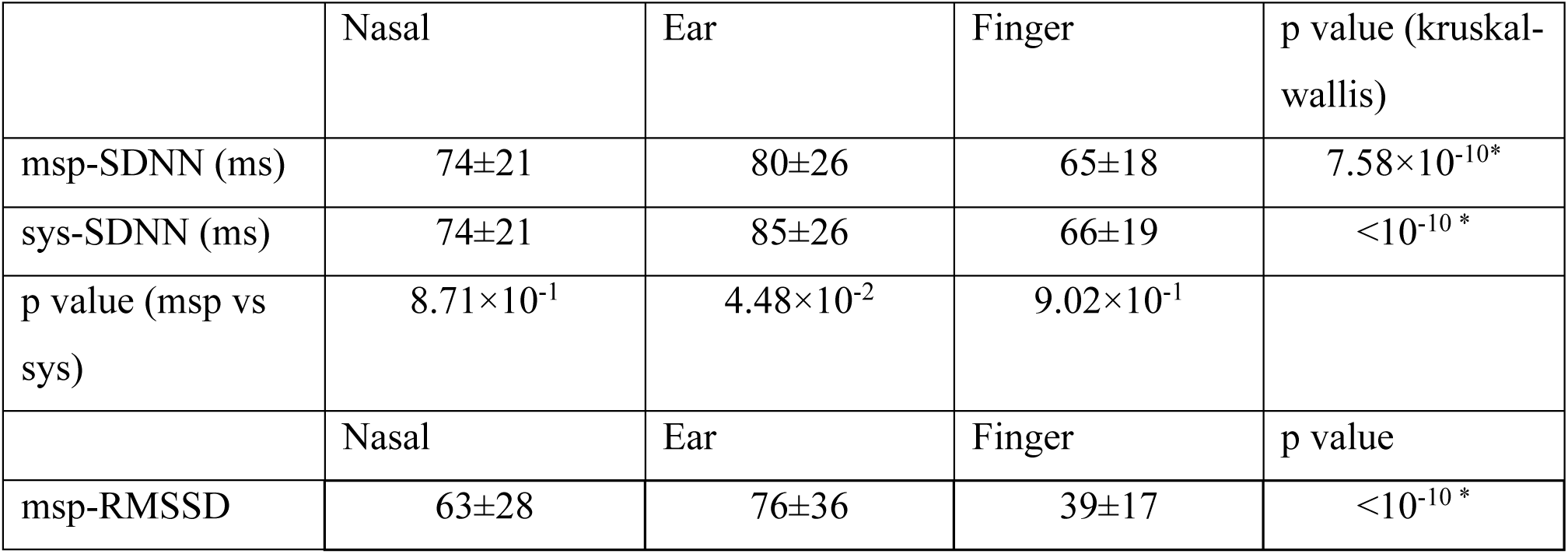
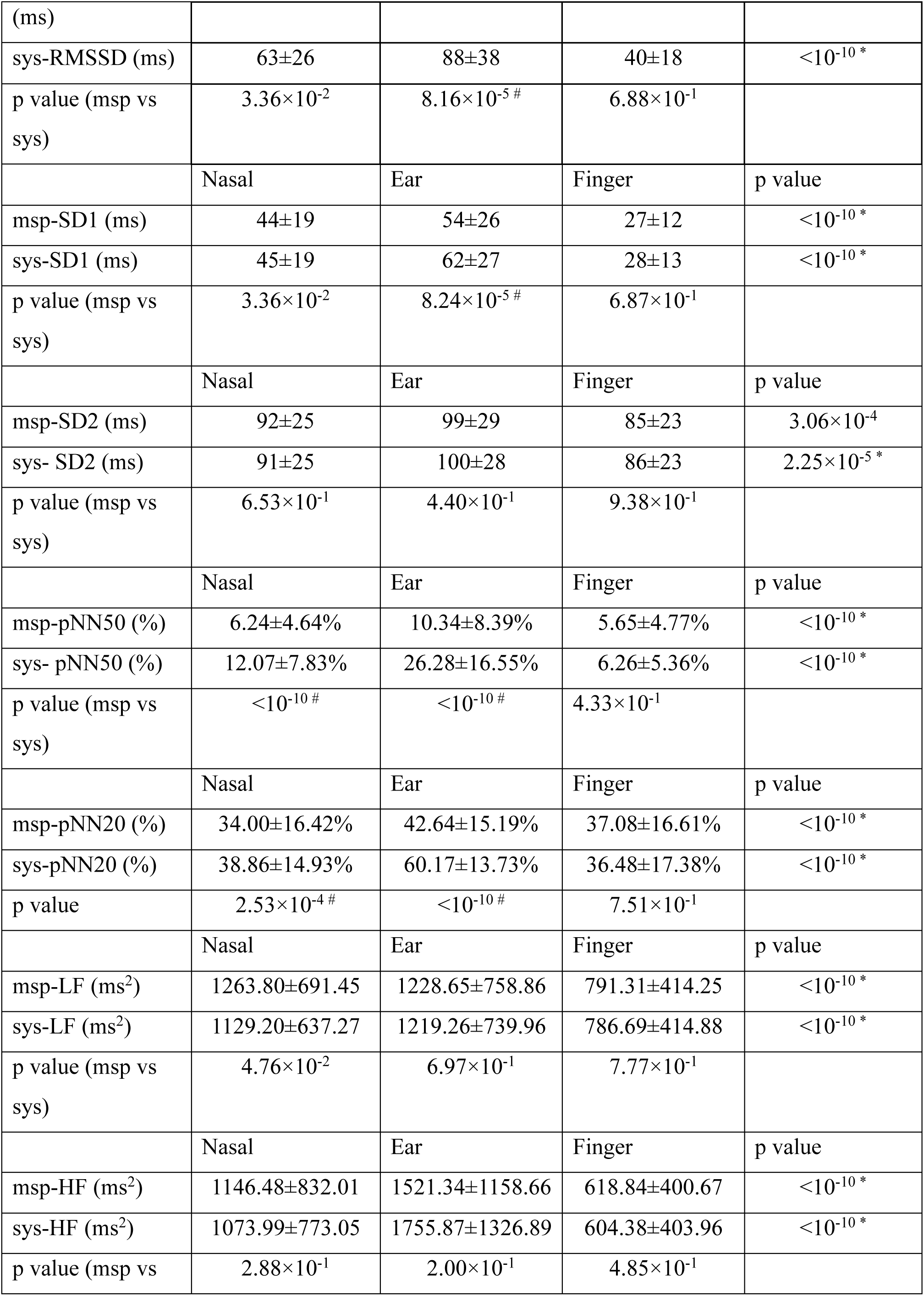
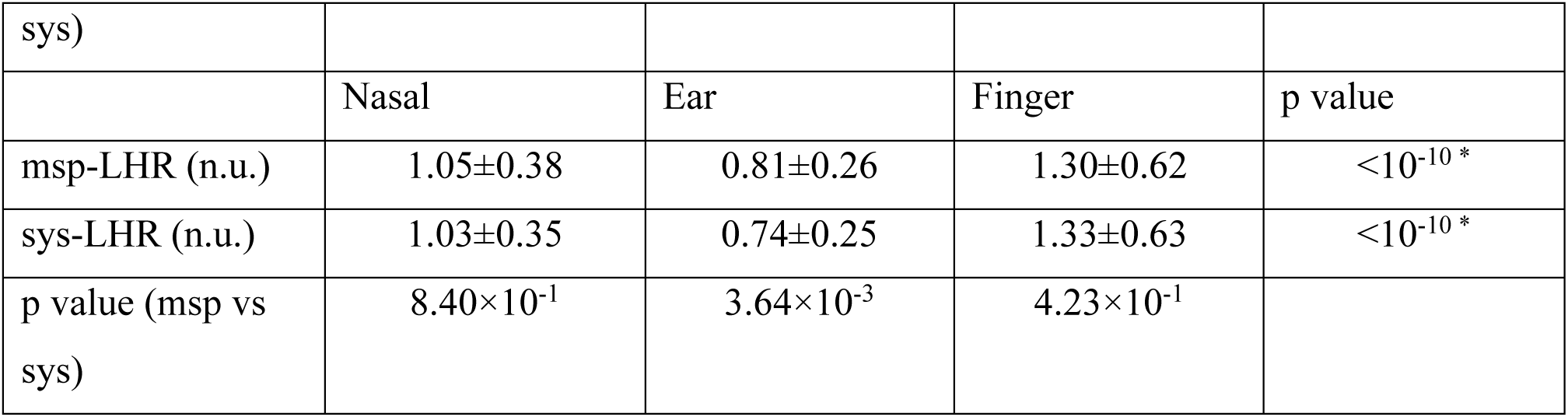
2. Table5 without SQI

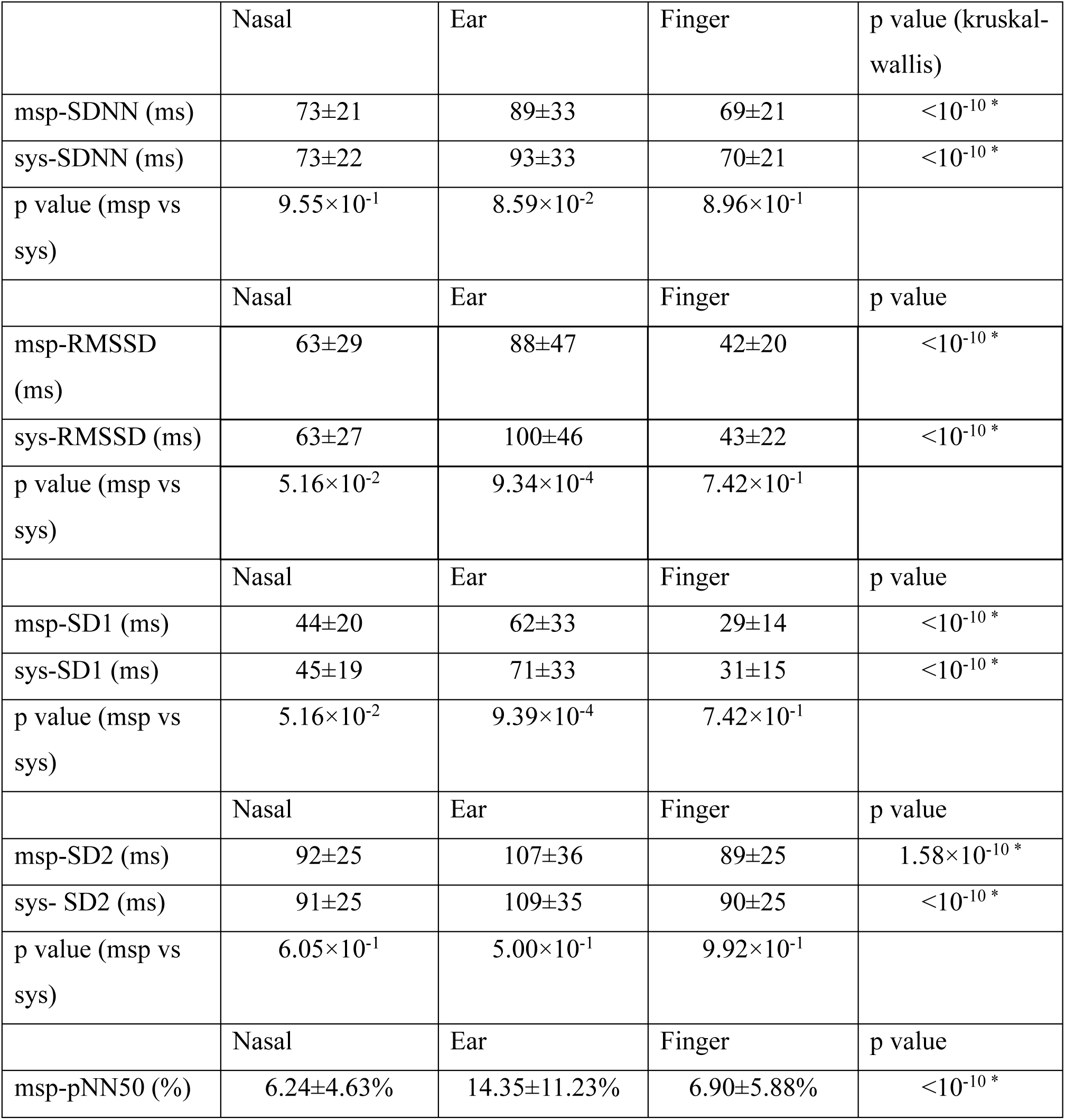
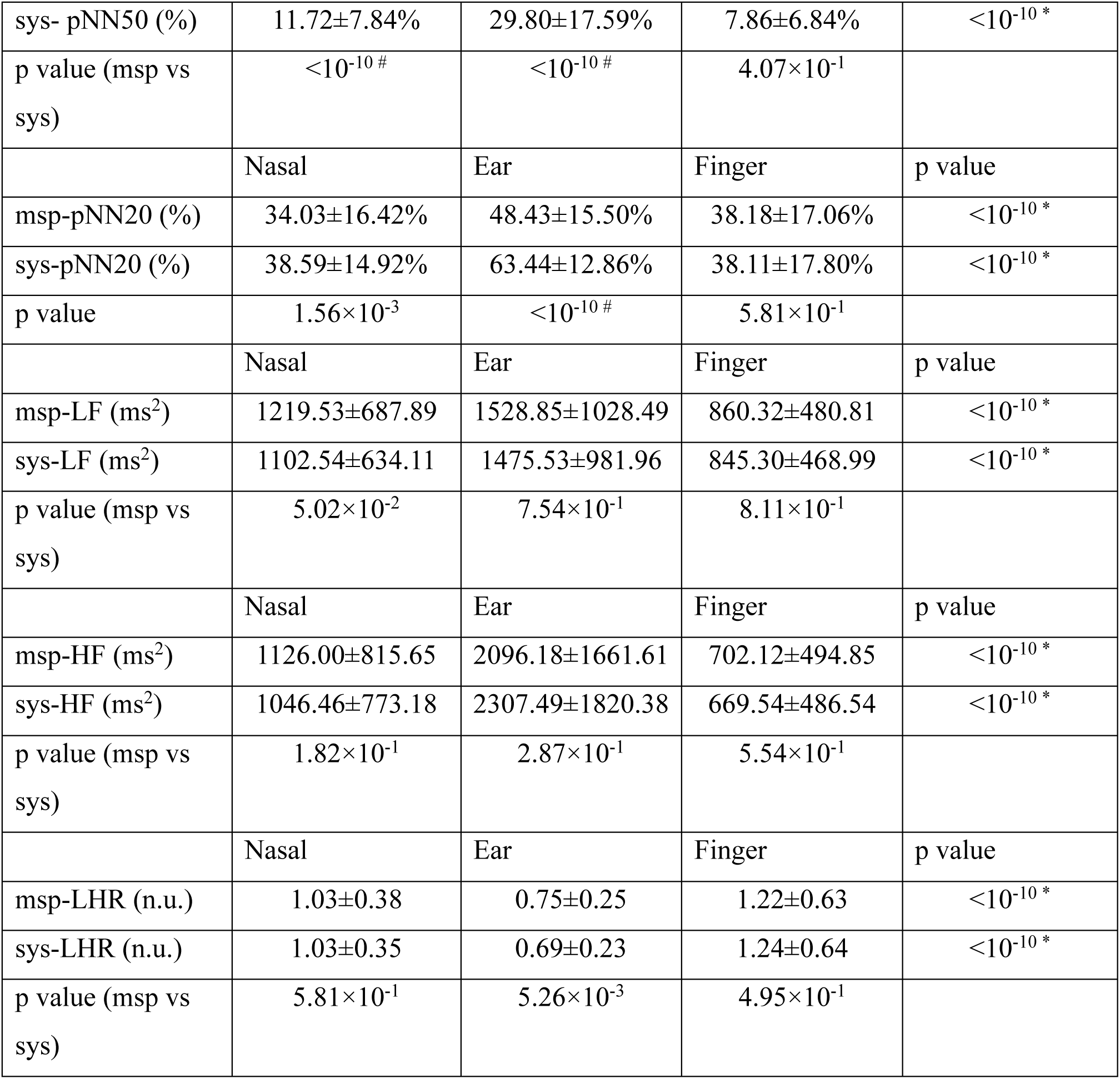

E. Table 6 with and without SQI

1. Table 6 with SQI

2. Table 6 without SQI

|  | Nasal | Ear | Finger | p value |
| --- | --- | --- | --- | --- |
| PHI2 (radian) | 1.77±0.20 | 2.16±0.29 | 1.36±0.30 | <10<sup>-10</sup> * |
| PHI3 (radian) | 1.80±1.34 | -2.07±0.54 | 2.22±0.37 | <10<sup>-10</sup> * |
| AHI2 (a.u) | 0.50±0.16 | 0.38±0.14 | 0.59±0.13 | <10<sup>-10</sup> * |
| AHI3 (a.u) | 0.26±0.06 | 0.19±0.06 | 0.25±0.06 | <10<sup>-10</sup> * |

|  | Nasal | Ear | Finger | p value |
| --- | --- | --- | --- | --- |
| PHI2 (radian) | 1.75±0.22 | 2.09±0.37 | 1.35±0.32 | <10<sup>-10</sup> * |
| PHI3 (radian) | 1.47±1.85 | -2.02±0.78 | 2.12±0.48 | <10<sup>-10</sup> * |
| AHI2 (a.u) | 0.49±0.17 | 0.38±0.15 | 0.56±0.15 | <10<sup>-10</sup> * |
| AHI3 (a.u) | 0.25±0.06 | 0.18±0.07 | 0.24±0.08 | <10<sup>-10</sup> * |

